# Evidence for patterns of colonization risk in the nationwide hospitalized patient network in France

**DOI:** 10.64898/2026.09.17.26363228

**Authors:** Shrichand Bhuria, Lucille Calmon, Madhav Chaturvedi, Laura Temime, André Karch, Tjibbe Donker, Vittoria Colizza, Pascal Crépey

## Abstract

Patient movement within and between hospitals creates a complex hospital network that facilitates the spread of antimicrobial-resistant bacteria and contributes to healthcare-associated infections. Yet most national-scale studies infer spread from aggregated hospital-to-hospital patient sharing, leaving patient-to-patient contact pathways and the role of routine patient mobility in shaping colonization risk poorly characterized. Using France’s 2023–2024 national hospital-discharge data, we reconstructed a temporal patient-based network of 10.4 million patients across 1,408 acute-care hospitals, comprising 326.6 million overnight ward co-location interactions. We simulated colonization over 365 days on the observed day-by-day temporal network using a stochastic susceptible–colonized–susceptible model, with a mean colonization duration of 180 days. We used national carbapenemase-producing Enterobacterales (CPE) surveillance data to calibrate differences in transmission risk across clinical settings. Across 2,000 simulations initiated on randomly selected dates in 2023, we calculated patient-level colonization probabilities and summarized risk across patients’ residential areas, wards, and hospitals. We further assessed hospital-level consistency between simulated risk and recorded CPE occurrence. Simulated risk was highly heterogeneous across patients and care settings. When mapped by place of residence, hospital-associated colonization risk showed strong spatial clustering in both urban and rural areas. Multivariable regression analyses identified patient- and hospital-level characteristics associated with heterogeneity in simulated colonization risk. Sensitivity analyses varying initial seeding and colonization duration showed that relative risk patterns remained highly stable. This nationwide framework shows how patient mobility and ward co-location patterns can reveal patterns of hospital-associated colonization risk and could support pathogen-specific surveillance and intervention evaluation.

**Significance Statement:** Asymptomatic carriage allows antimicrobial-resistant bacteria to spread through healthcare systems while colonized patients may remain unrecognized in the absence of active screening. We reconstructed a nationwide, time-resolved network of ward co-location among hospitalized patients in France. We accounted for differences in transmission risk across clinical settings using national surveillance data on carbapenemase-producing Enterobacterales. Using a transmission model, we characterized colonization-risk patterns and identified patient- and hospital-level features associated with variation in risk. This framework offers health authorities a way to examine where patient movement and hospital care may contribute to concentrations of colonization risk, although pathogen- and context-specific adaptation and evaluation remain necessary to guide surveillance or interventions.

**Author contributions:** S.B., L.C., L.T., A.K., T.D., V.C., and P.C. designed research; S.B. and P.C. performed research; S.B. and P.C. analyzed data; S.B. and P.C. wrote the paper; all authors reviewed and edited the final manuscript.

## Introduction

Antimicrobial resistance (AMR) is widely recognized as a major global health threat: in 2019, drug-resistant infections were directly responsible for an estimated 1.27 million deaths and were associated with nearly 5 million deaths worldwide [1, 2]. Antibiotic-resistant bacteria (ARB) are particularly concerning because they can resist commonly used antibiotics and may colonize individuals for prolonged periods without causing symptoms; such asymptomatic carriage may remain unrecognized in the absence of active screening [3, 4]. During asymptomatic carriage, colonized individuals can transmit resistant bacteria in both community and healthcare settings through close contact and contaminated environments [5–7]. In healthcare settings, prolonged hospitalization, invasive procedures, and intensive antibiotic exposure create substantial opportunities for ARB colonization and infection [8, 9]. Healthcare-associated infections (HAIs)—infections acquired during hospitalization—remain common: the World Health Organization estimates that about 7% of hospitalized patients in high-income countries and 15% in low- and middle-income countries acquire at least one HAI during their stay, many of which are caused by ARB [10]. Infections caused by ARB are often harder to treat and are associated with increased morbidity, prolonged hospital stays, higher mortality, and greater healthcare costs [2, 11]. In turn, HAIs increase antibiotic use, which promotes the selection and spread of resistance, making HAIs increasingly difficult to control and creating a self-reinforcing feedback loop between AMR and HAIs [12–14].

Hospitals are not only sites of care but also key drivers, reservoirs, and amplifiers of ARB transmission, as healthcare infrastructure, ward environments, and patient pathways can promote and sustain the spread of resistance through direct and indirect mechanisms [15]. At the ward level, patients share confined spaces, are indirectly connected through healthcare personnel who move between rooms, and are exposed to shared equipment and surfaces, creating repeated opportunities for ARB acquisition and onward transmission. These opportunities are especially important among medically fragile individuals undergoing invasive procedures (e.g., surgery, intravenous lines, and urinary catheters), which increase susceptibility to colonization and infection [16–19]. Importantly, transmission is not confined to a single hospital. Patients move between hospitals and other healthcare facilities for follow-up care, referrals, and readmissions, thereby linking healthcare facilities over time and creating nationwide pathways for dissemination [20–22]. As a result, ARB can spread within and between healthcare facilities through the movement of asymptomatically colonized patients. After discharge, colonized patients may carry ARB back into the community; conversely, individuals colonized in the community may reintroduce ARB into healthcare facilities upon admission or readmission [23, 24]. This bidirectional flow blurs the line between healthcare-associated and community-associated transmission, complicating both surveillance and control strategies. Overall, understanding how ARB disseminates through a complex healthcare system requires considering patient mobility: when and where patients receive care, whom they share space with, and where they return after discharge.

Antibiotic-resistant bacteria do not spread in isolation; rather, they emerge, persist, and circulate through the coupled dynamics of patient contacts, healthcare environments, and mobility across care settings [25]. To better understand these dynamics, researchers have long used data-driven mathematical models and network-based approaches to study the transmission of ARB within healthcare systems [26–33]. In recent years, well-established national hospital databases from the United States [34–37] and Europe—including England, France, Germany, and the Netherlands [20–22, 38–43]—have enabled the reconstruction of large-scale healthcare networks in which nodes are hospitals and edges represent direct patient transfers or indirect links through sequential admissions between hospitals. These hospital-level models have been instrumental for simulating regional spread, estimating epidemic risk, and informing coordination of infection prevention. However, they obscure the fine-scale contact structure that drives transmission—specifically, who is in contact with whom, where, and when. A handful of recent studies have demonstrated the value of such individual-level networks built from within-hospital patient co-location data, but most are limited to single hospitals or regions or to specific patient groups, restricting their generalizability and national relevance [44–50]. As a result, patient-to-patient contact pathways—and the structural component of colonization risk induced by routine patient mobility—remain poorly characterized at the national scale.

To overcome these limitations, we reconstructed a data-driven, individual-level contact network from hospitalization records covering patients admitted to public and private acute-care hospitals throughout metropolitan France. These records came from France’s national hospital discharge database (French acronym: PMSI; *Programme de Médicalisation des Systèmes d’Information*) [51], specifically its *Médecine, Chirurgie, Obstétrique* extract (PMSI–MCO), which covers acute-care stays in medicine, surgery, and obstetrics. PMSI–MCO records admission and discharge dates and ward-level movements, allowing individual hospital trajectories to be reconstructed over time. Using these trajectories, we identified when and where patients were co-located and reconstructed potential patient-to-patient transmission opportunities on a day-by-day basis, capturing the fine-scale temporal organization of patient movement within and between wards and hospitals at the national scale. Ward co-location does not necessarily imply direct physical contact, but represents a plausible opportunity for ARB transmission through patient proximity or indirect exposure mediated by healthcare personnel, shared equipment, or the ward environment. Long-term care facilities, rehabilitation centers, and other settings outside the acute-care sector were not represented, potentially omitting additional transmission pathways within the broader healthcare system.

Using this high-resolution contact structure, we developed a susceptible–colonized–susceptible model allowing colonization clearance and recolonization and used it to characterize colonization risk over a 365-day period following stochastic introduction into the observed day-by-day temporal network. To represent heterogeneity in transmission risk across clinical settings, we used carbapenemase-producing Enterobacterales (CPE) reports from e-SIN (*signalement externe des infections nosocomiales*), the French national electronic reporting system for healthcare-associated infections [52], to calibrate relative transmission risk across 12 major clinical groups of wards. From repeated simulations, we derived patient-level colonization probabilities. We further used surveillance data to assess post-calibration consistency between simulated and surveillance-derived patterns at the clinical-group level and separately examined whether hospital-level simulated risk was associated with recorded CPE occurrence. Within this framework, we addressed three related questions: (i) how simulated patient colonization risk was spatially distributed across France when aggregated by patients’ places of residence and by their points of care (hospitals and ward types); (ii) whether these simulated patterns were consistent with those observed in national CPE surveillance; and (iii) which patient- and hospital-level characteristics were most strongly associated with elevated simulated colonization risk. We addressed these questions using national risk mapping, spatial analyses, surveillance comparisons, and multivariable beta-regression models. Full methodological details are provided in the *Materials and Methods* section.

## Results

Using nationwide hospital discharge data from France in 2023–2024, we reconstructed a day-by-day patient-based network (PBN) capturing ward-level overnight co-location among hospitalized patients. We then simulated colonization spread on this day-by-day temporal network using a stochastic susceptible–colonized–susceptible model allowing clearance and recolonization. To represent heterogeneity in transmission risk across clinical settings, we calibrated relative transmission multipliers for 12 major clinical groups using national CPE surveillance data, so that the relative distribution of simulated colonizations across clinical groups reflected the surveillance-derived profile after accounting for inpatient exposure. We performed 2,000 simulations over 365 days, each initiated on a randomly selected date in 2023 with 10 randomly selected colonized patients; the primary analysis assumed a mean colonization duration of 180 days. For each patient, colonization probability was defined as the proportion of simulation runs in which the patient became colonized along their observed hospital trajectory. These patient-level probabilities were then aggregated at different levels to characterize simulated colonization risk across residential areas, clinical groups, ward types, and hospitals. Because patients could become colonized in different clinical settings across simulation runs, hospital-, ward-type-, and clinical-group-level risks were based on the corresponding setting-attributed patient probabilities. Residential-area risk instead used each patient’s overall colonization probability, mapped according to place of residence. Detailed definitions and aggregation procedures are provided in the *Materials and Methods* section.

### Patient-level colonization risk mapping across metropolitan France

To examine how simulated hospital-associated colonization risk was distributed across patients’ places of residence, we constructed a bivariate Voronoi map of residential colonization risk and patient volume across metropolitan France (Fig. 1A). Metropolitan France was partitioned into 5,533 Voronoi cells, each corresponding to a unique valid PMSI residential geographic code and the PBN patients residing in that area. For each area, the residential colonization risk ratio was defined as the mean simulated colonization probability among patients living in that area divided by the national mean simulated colonization probability across all PBN patients. Patient volume was expressed relative to the national mean number of PBN patients across residential areas. The bivariate map revealed marked spatial heterogeneity, with elevated colonization risk occurring in areas with both high and low patient volumes, indicating that the spatial concentration of simulated risk was not explained by patient volume alone.

**Figure 1:**
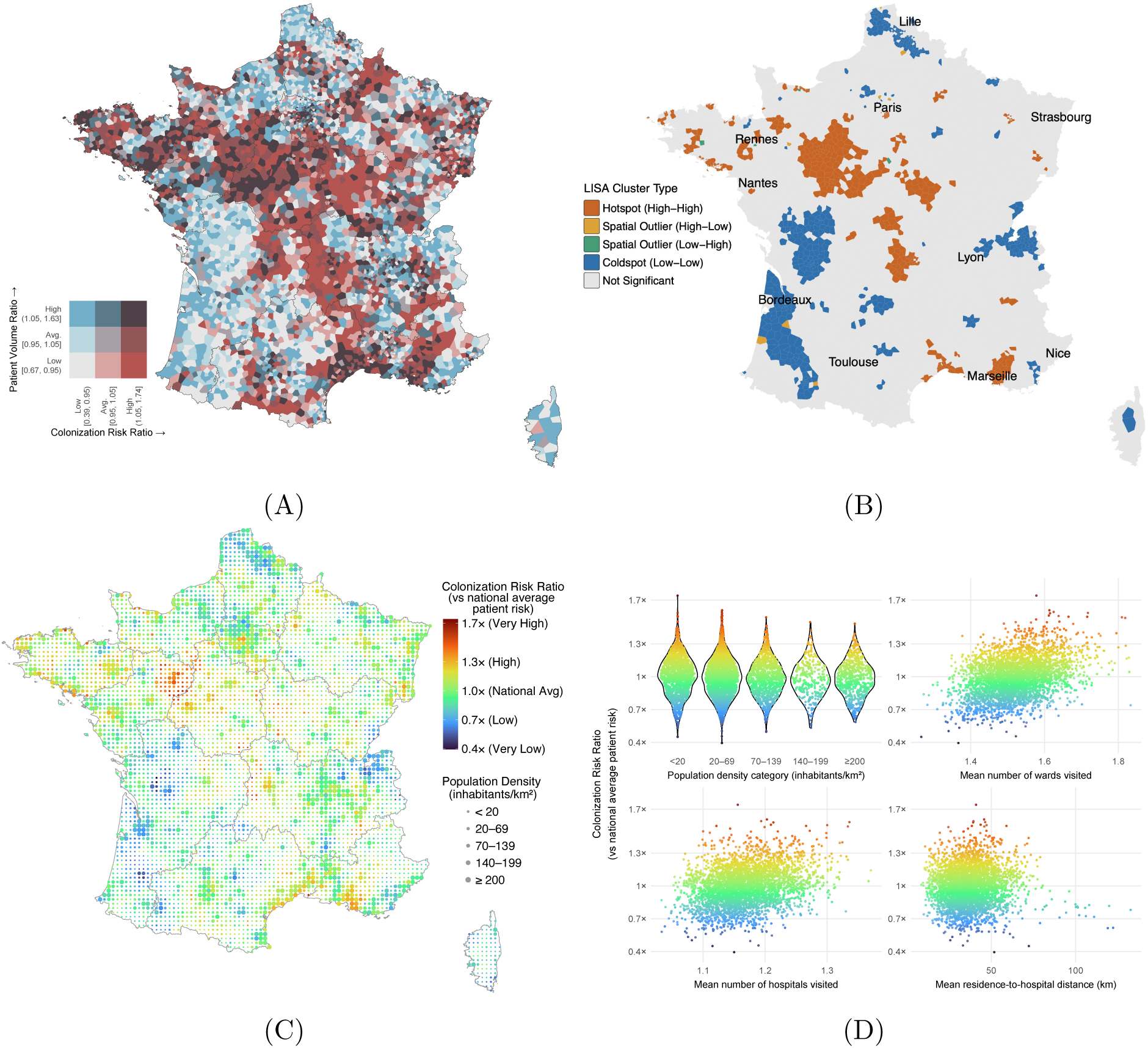
Spatial distribution and descriptive correlates of simulated patient-level colonization risk in France. **(A)** Bivariate Voronoi map of metropolitan France partitioned into 5,533 residential cells, each corresponding to a unique valid PMSI residential geographic code and the PBN patients residing in that area. Polygon color encodes two quantities: the residential colonization risk ratio (horizontal scale), defined as the mean simulated colonization probability among patients living in that area relative to the national mean across all PBN patients, and the patient volume ratio (vertical scale), defined as the number of PBN patients residing in that area relative to the national mean across residential areas. **(B)** Local Indicators of Spatial Association (LISA) map showing high–high hotspots, low–low coldspots, and high–low or low–high spatial outliers in residential colonization risk. **(C)** Dot-density map of metropolitan France in which each dot represents a population-density grid cell; dot size indicates the population-density category, and dot color indicates the residential colonization risk ratio relative to the national mean simulated patient risk. **(D)** Descriptive associations between residential colonization risk and area-level characteristics. The violin plot shows the distribution of residential colonization risk ratios across population-density categories. The scatter plots show how residential colonization risk ratio varies with the mean number of wards visited per patient, the mean number of hospitals visited per patient, and the mean residence-to-hospital travel distance among patients living in each residential area. Each point corresponds to one dot in panel **C**.

To assess whether the geographic pattern in Fig. 1A was spatially structured, we quantified spatial autocorrelation in residential colonization risk ratios. Global Moran’s *I* was 0.66 (95% CI: 0.64–0.67, *P <* 0.001), indicating positive spatial autocorrelation. To localize this structure, we applied Local Moran’s *I*, a local indicator of spatial association (LISA), which identified high–high hotspots and low–low coldspots, with relatively few high–low or low–high spatial outliers (Fig. 1B). Together, these results indicate a clear, non-random geographic structure in simulated residential colonization risk. Full details of the spatial-autocorrelation analysis are provided in the *Materials and Methods* and the *SI Appendix*.

While the analyses above identify *where* residential colonization risk concentrates, we next explored selected area-level characteristics associated with these spatial patterns. A natural first hypothesis was that risk simply reflected population density. To explore this, Fig. 1C overlays residential colonization risk ratios on population-density categories (GHSL data) using a regular dot grid across metropolitan France. The resulting map shows that elevated risk is not confined to major metropolitan centers: many sparsely populated rural areas exhibit above-average risk, whereas several high-density urban zones show comparatively low values. We next explored how residential colonization risk varied with patients’ use of the hospital system and with residence-to-hospital travel distance, used here as a simple measure of the geographic separation between patients’ places of residence and the hospitals they attended (Fig. 1D). The violin plot shows substantial overlap in risk across population-density categories, indicating that population density alone did not account for the observed spatial heterogeneity. The scatter plots show that residential colonization risk tended to increase across the observed ranges of both the mean number of wards visited and the mean number of hospitals visited by patients living in each residential area. These measures were calculated at the patient level and then averaged within residential areas. Mean residence-to-hospital travel distance was calculated for each patient as the straight-line distance between the residential location and the hospitals visited, weighted by the number of admissions to each hospital when multiple hospitals were visited, and then averaged within residential areas. In contrast to the patterns observed for ward and hospital use, residential colonization risk showed no clear monotonic pattern across mean travel distance. Residential-area summaries were assigned to the population-density grid cells shown in Fig. 1C and, where multiple residential areas contributed to a grid cell, combined using the number of PBN patients as weights. Together, these descriptive analyses indicate that residential colonization risk was not simply patterned by population density or residence-to-hospital distance and also varied with patients’ patterns of hospital use. Additional descriptive analyses of continuous population density (scatter plot), hospital admissions per 1,000 residents, mean patient degree, mean length of stay per ward, and residential socioeconomic deprivation (FDep20; French Deprivation Index) are provided in the *SI Appendix* (Fig. S4) as complementary analyses beyond the principal residential characteristics shown in Fig. 1D.

### Colonization risk across clinical groups

To examine how simulated patient-level colonization risk varied across clinical care settings, we summarized colonization probabilities attributed to the detailed ward types recorded in the PMSI database and mapped these ward types to the 12 broader clinical groups used in the transmission model (Fig. 2; see *Materials and Methods*). These groups combined clinically similar ward types while allowing a consistent mapping between PMSI and the clinical-service categories available in the national e-SIN surveillance data used for calibration. Full ward-type-specific results are provided in the *SI Appendix* (Fig. S7), and the mapping of ward types to clinical groups is provided in Table S4. Patient-level colonization risk ratios varied substantially both between and within clinical groups (Fig. 2A). The mean colonization risk ratio was highest in oncology, adult intensive care, cardiology, adult medicine, and pediatric intensive care, whereas obstetrics/gynecology, emergency/short-stay, and palliative or specialized care had the lowest mean values. The broad within-group distributions further showed that patients receiving care within the same clinical group could have markedly different simulated risks, consistent with differences in their hospital trajectories and ward co-location histories. We then assessed post-calibration consistency with surveillance by comparing the simulated clinical-group profile with the corresponding 2023 e-SIN surveillance profile after standardizing both for inpatient exposure, measured as the total number of patient-days in each clinical group (Fig. 2B). This standardization identified groups accounting for more or fewer simulated colonizations or reported CPE cases than expected from their volume of inpatient activity. The simulated and surveillance-derived profiles were strongly rank-concordant (Spearman *ρ* = 0.97): groups with relatively high simulated colonization rates generally also had relatively high surveillance-derived relative reported-case rates, whereas groups with lower simulated rates tended to remain below the national reference in surveillance. Several groups nevertheless departed from the line of equality, indicating that agreement was stronger for the relative ordering of clinical settings than for the exact magnitude of their rates. Because pooled e-SIN data from 2017–2025 were used to calibrate the clinical-group transmission multipliers, this comparison represents post-calibration empirical consistency rather than an independent validation.

**Figure 2:**
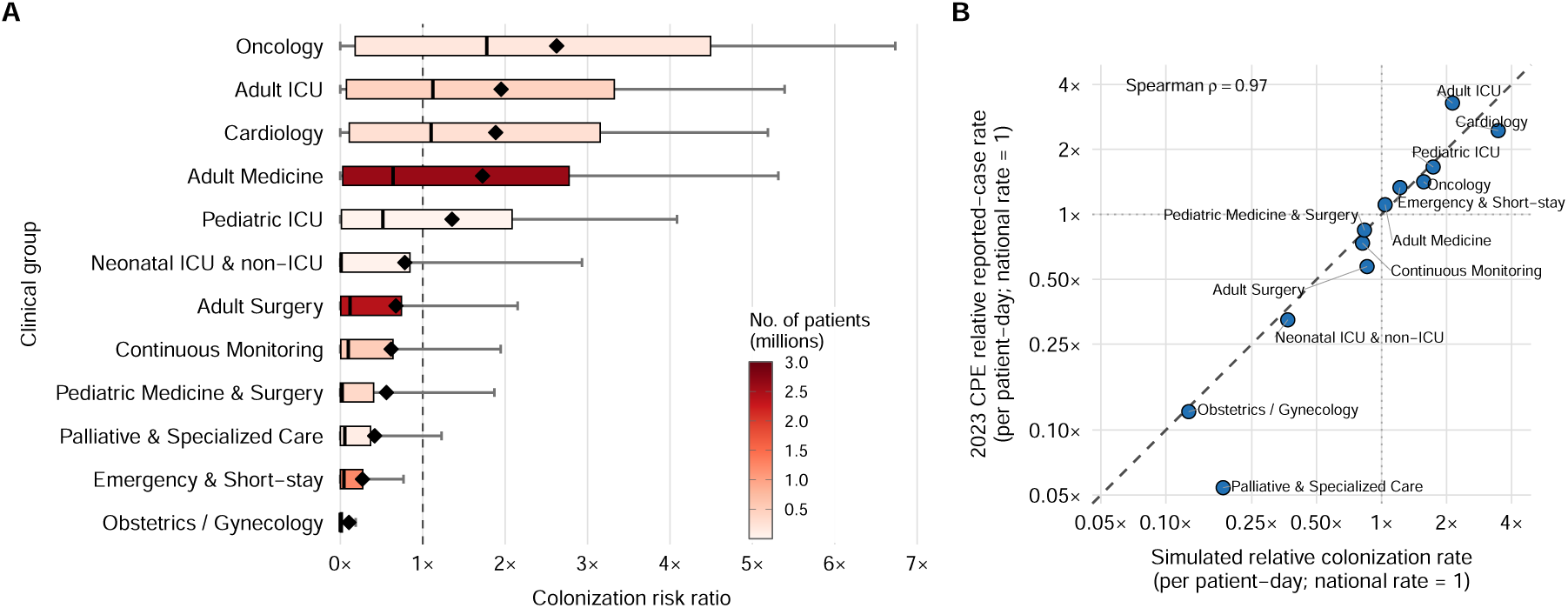
Clinical-group heterogeneity in simulated colonization risk and concordance with 2023 CPE surveillance. **(A)** Distribution of simulated patient-level colonization risk ratios across the 12 clinical groups. Within each clinical group, patient-level colonization risk ratios were calculated by dividing each patient’s colonization probability attributed to that group by the national mean across all patient–clinical-group observations. The dashed vertical line marks the national reference value of 1. Gray whiskers show the 10th–90th percentiles, boxes show the interquartile range, black vertical lines indicate the median, and black diamonds indicate the mean. Box color represents the number of patients with at least one stay in each clinical group. **(B)** Concordance between simulated and surveillance-derived clinical-group profiles after standardization for inpatient exposure. For each clinical group, expected simulated colonizations were obtained by summing across patients their colonization probabilities attributed to that clinical group, estimated from the 2,000 primary simulations. The simulated relative colonization rate was then calculated as the clinical group’s share of the total expected simulated burden divided by its share of national inpatient exposure in 2023. Inpatient exposure was measured using patient-days, defined as the total number of days spent by patients in each clinical group. The surveillance relative reported-case rate was calculated analogously as the clinical group’s share of weighted CPE cases reported through e-SIN in 2023 divided by its share of national patient-days. A value of 1 indicates that the group’s share of simulated colonizations or reported cases was proportional to its share of national inpatient exposure, whereas values above or below 1 indicate higher or lower relative rates, respectively. Each point represents one clinical group. The dashed diagonal indicates equality between the simulated and surveillance-derived relative rates, whereas the dotted horizontal and vertical lines mark the national reference value of 1. Both axes are shown on logarithmic scales.

### Hospital-level colonization risk is heterogeneous across France but only weakly spatially structured

To examine how simulated colonization risk was distributed across hospitals, we averaged patient-level hospital-attributed colonization probabilities within each hospital. For visualization, we then grouped nearby hospitals into spatial clusters and summarized each cluster using a weighted mean hospital-level risk, with the number of PBN patients in each hospital used as weights (Fig. 3A). This procedure improved map readability and allowed nearby hospitals that may share catchment areas or referral pathways to be summarized jointly without imposing administrative boundaries. The hospital-level colonization risk ratio therefore indicates whether the mean hospital-attributed colonization risk among patients associated with hospitals in a given polygon is above or below the national mean hospital-attributed patient colonization risk across France (*Materials and Methods*). The resulting map showed marked heterogeneity across the hospital network, with clusters of relatively high and low colonization risk interspersed throughout metropolitan France and no clear large-scale geographic gradient. Figure 3B further shows descriptive relationships between cluster-level colonization risk and hospital capacity, annual admissions, mean length of stay, and the proportion of patients with repeated admissions. High- and low-risk clusters occurred across a broad range of these characteristics, although higher-risk clusters tended to be more frequent among settings with greater cumulative patient exposure and healthcare use. Substantial variation nevertheless remained within each structural characteristic, indicating that no single measure alone captured the observed heterogeneity.

**Figure 3:**
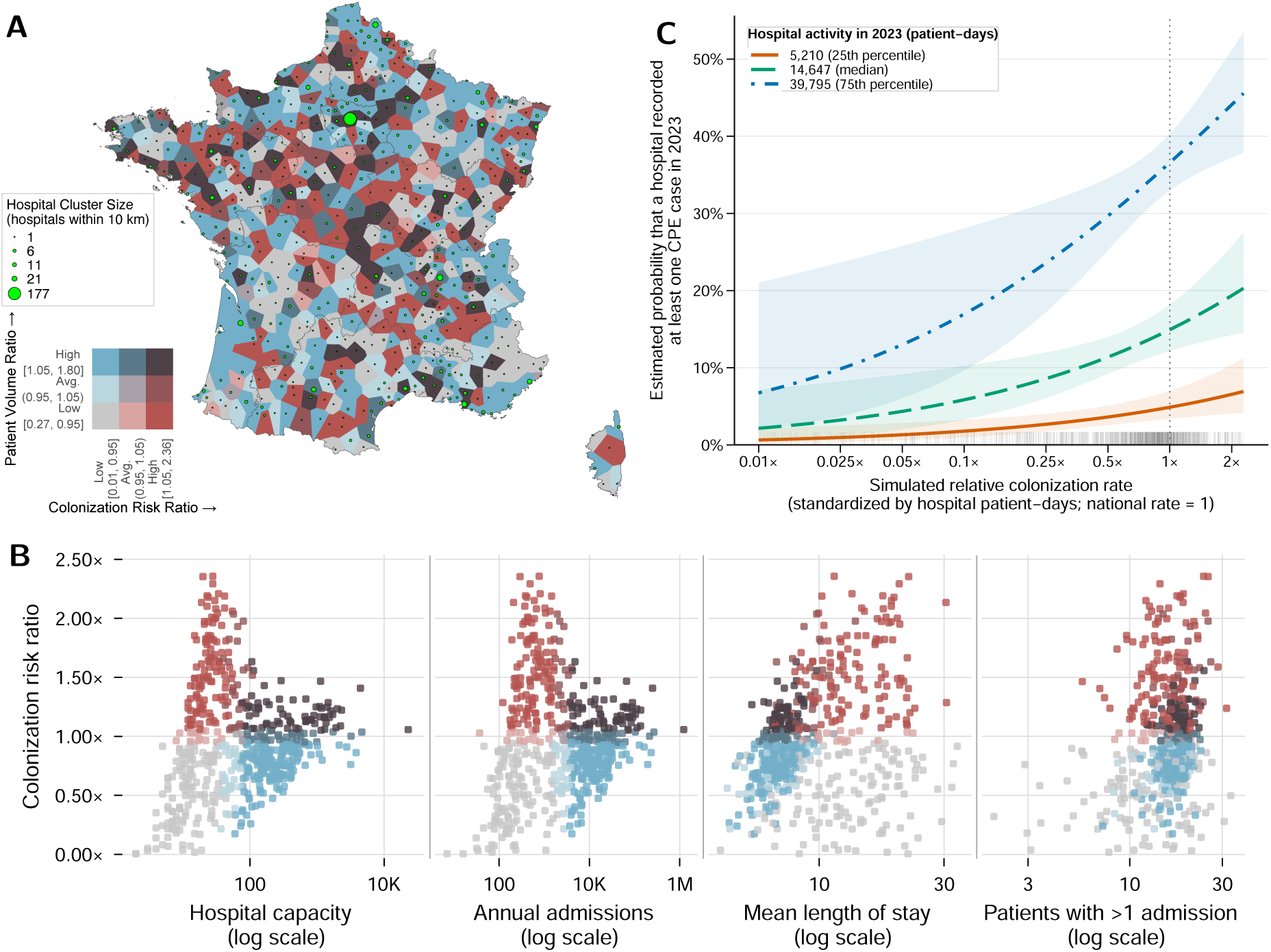
Hospital-level simulated colonization risk, hospital characteristics, and consistency with 2023 CPE surveillance. **(A)** Bivariate Voronoi map of metropolitan France in which each polygon represents a spatial cluster of hospitals located within 10 km of each other. Polygon color encodes the hospital-level colonization risk ratio (horizontal gradient), defined as the mean hospital-attributed colonization risk among patients who visited hospitals in that polygon relative to the national mean patient-level hospital-attributed colonization risk. The vertical gradient shows the patient volume ratio, defined relative to the national mean. Green point size indicates the number of hospitals contained in each polygon. **(B)** Descriptive associations between polygon-level colonization risk ratio and four structural characteristics calculated at the same polygon level: hospital capacity, total annual admissions, mean length of stay, and the proportion of patients with more than one admission. Each point represents one hospital cluster and is colored according to the same 3×3 risk–volume classification shown in panel (A). **(C)** Association between simulated relative colonization rate and the probability that a hospital recorded at least one CPE case in 2023. Patient-level hospital-attributed colonization probabilities from the 2,000 primary simulations were summed within each hospital to obtain its expected simulated colonization burden. To account for differences in hospital activity, the simulated relative colonization rate was defined as the hospital’s share of the total expected simulated burden divided by its share of national inpatient exposure in 2023, measured in patient-days (total days spent by patients in the hospital). A value of 1 therefore indicates that the hospital’s share of simulated colonizations was proportional to its share of national inpatient exposure. Curves show adjusted probabilities from a multivariable logistic regression in which the binary outcome was whether the hospital recorded at least one CPE case in 2023. The predictors were simulated relative colonization rate and hospital patient-days, both entered as continuous log_2_-transformed variables; thus, a one-unit increase corresponds to a doubling of the original value. Predictions are shown for hospitals at the 25th percentile, median, and 75th percentile of patient-days, while patient-days remained continuous in the fitted model. Shaded areas show 95% confidence intervals, the vertical dotted line marks the national reference value of 1, and rug marks show the observed distribution of simulated relative colonization rates.

To quantify the geographic structure of hospital-associated colonization risk, we assessed spatial autocorrelation across hospital Voronoi polygons. Global Moran’s *I* was 0.095 (approximate 95% CI: 0.046–0.144; *P <* 0.001), indicating very weak positive spatial autocorrelation. At the local level, Local Moran’s *I* (LISA) identified no high–high hotspots, low–low coldspots, or high–low or low–high spatial outliers after adjustment for multiple testing. Together, these results show that hospital-associated colonization risk was highly heterogeneous but only weakly geographically clustered, in contrast to the much stronger spatial structure observed when patient risk was mapped by place of residence (Global Moran’s *I* =0.66). Additional methodological details, including the Moran scatterplot, are provided in the *Materials and Methods* and the *SI Appendix* (Fig. S8).

### Hospital-level consistency with CPE surveillance

We next examined whether hospital-level variation in simulated relative colonization rate was associated with CPE occurrence reported through e-SIN in 2023. Among the 1,391 hospitals with both simulated risk estimates and 2023 e-SIN surveillance data, only 312 (22.4%) recorded at least one CPE case, whereas the remaining hospitals had no recorded case. Given this predominance of zero reported cases, we used a binary outcome indicating whether each hospital recorded at least one case during the year rather than modeling case counts. We used a multivariable logistic regression to assess whether simulated relative colonization rate was associated with recorded CPE occurrence after accounting for hospital activity, measured using hospital patient-days (the total number of days spent by patients in each hospital in 2023). As shown in Fig. 3C, the estimated probability that a hospital recorded at least one CPE case increased with its simulated relative colonization rate across different levels of hospital activity. After adjustment for patient-days, each doubling of the simulated relative colonization rate was associated with 37% higher odds of recording at least one CPE case (adjusted odds ratio = 1.37, 95% CI: 1.13–1.69, *P* = 0.002). Adding the simulated relative colonization rate to a model containing patient-days alone also improved model fit (likelihood-ratio *χ*^2^ = 11.07, *P <* 0.001). These findings indicate that the simulated hospital-level risk measure contained information associated with recorded CPE occurrence beyond hospital activity alone. Further details on hospital linkage, regression specification, and model-based predictions are provided in the *SI Appendix*, Section S1.5.

### Patient- and hospital-level characteristics associated with simulated colonization risk

The risk maps and descriptive scatter plots above show substantial heterogeneity in simulated colonization risk across France. To identify patient- and hospital-level characteristics associated with this variation, we fitted separate multivariable beta-regression models (Fig. 4). Adjusted associations are presented as standardized risk ratios (SRRs), with values above or below 1 indicating higher or lower model-adjusted predicted colonization risk than the overall average, respectively. Full details of the modeling framework are provided in the *Materials and Methods* section. At the patient level (Fig. 4A), the clearest gradients were associated with the frequency, duration, and breadth of hospital exposure. Modeled risk increased progressively with the number of admissions, mean length of stay per ward, and number of wards visited, with the highest categories reaching approximately 3.1, 2.7, and 1.9 times the model-average risk, respectively. Age showed only modest variation, with SRRs remaining close to the model average across age groups (∼ 0.87–1.06). Residential socioeconomic deprivation, measured using the French Deprivation Index (FDep20; higher values indicate greater deprivation), likewise showed comparatively small differences around the model average. In contrast, patients who visited multiple hospitals had lower adjusted SRRs than those who visited a single hospital. This inverse association may reflect differences in the structure of healthcare exposure, as care distributed across several hospitals does not necessarily correspond to longer or more sustained ward-level exposure. Predominant ward type showed substantial heterogeneity in modeled risk. Higher-than-average SRRs were observed for several ward types within cardiology (15E, 02E, 02A), pediatric ICU (13G, 15H, 15C, 15I, 15G, 15D), adult ICU (02H, 02I, 02C, 02F, 01A, 02B, 01B), and oncology (40, 41). In contrast, lower-than-average SRRs were observed for ward types within obstetrics/gynecology (73, 71, 70), palliative and specialized care (08, 19, 20, 30), and emergency and short-stay care (07A, 07B). Estimates were less precise for some rare ward types, consistent with the smaller numbers of patients in these categories. Complete ward-type definitions and their broader clinical groupings are provided in the SI Appendix (Fig. S7 and Table S4), and complete patient-level model estimates are reported in Table S5.

**Figure 4:**
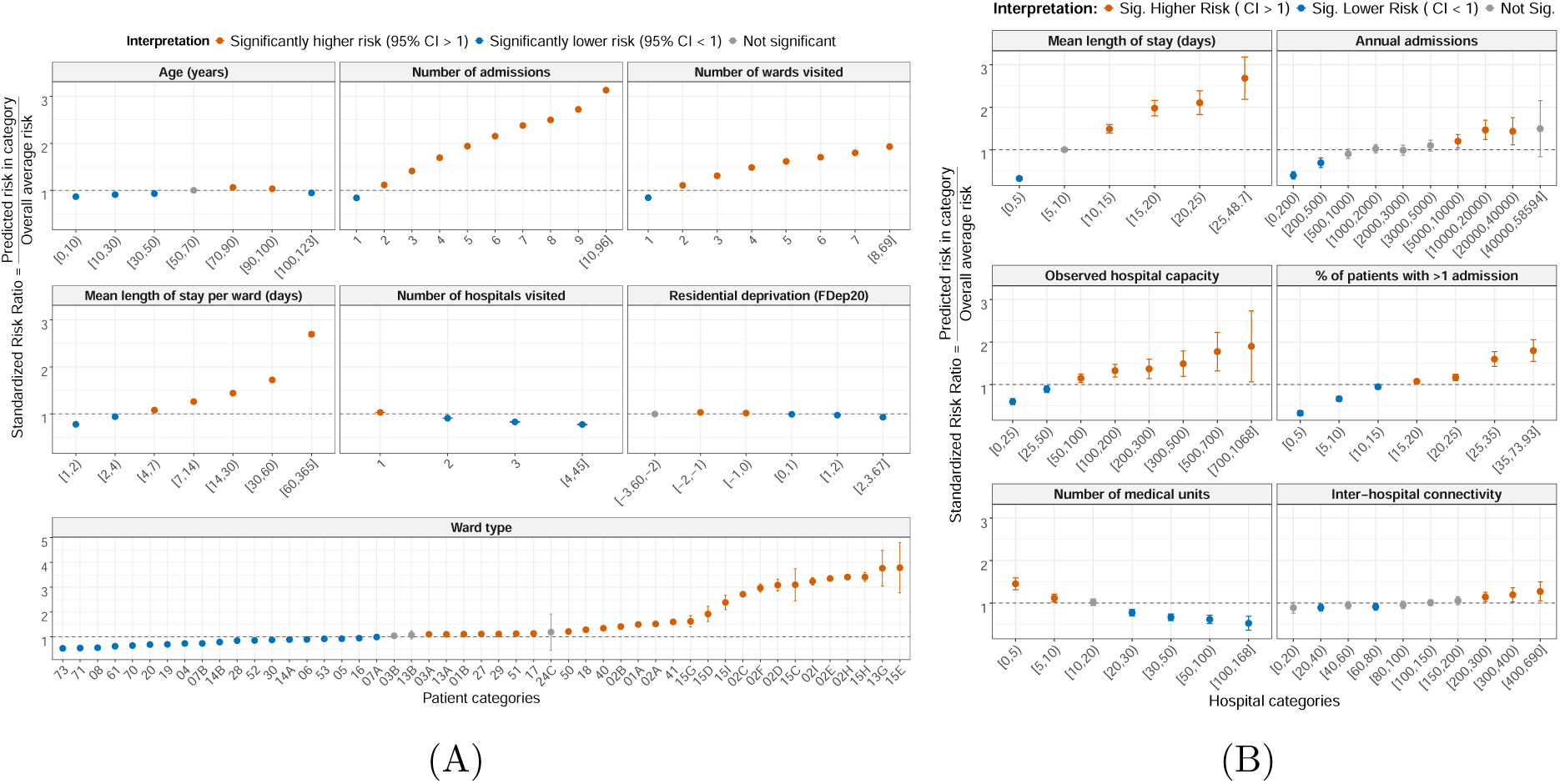
Patient- and hospital-level characteristics associated with simulated colonization risk. Separate multivariable beta-regression models were fitted at the patient and hospital levels. Adjusted associations are presented as standardized risk ratios (SRRs) derived from marginally standardized model predictions. For each predictor category, the SRR compares the adjusted predicted colonization risk with the overall average predicted risk from the corresponding model. Thus, SRR = 1 represents the model-specific average risk (horizontal dashed line). SRRs are interpreted within the patient- and hospital-level models separately. Points show SRRs with 95% confidence intervals. Orange and blue indicate intervals entirely above and below 1, respectively, and gray indicates intervals overlapping 1. **(A)** Selected patient-level characteristics: age, number of admissions, number of wards visited, mean length of stay per ward, number of hospitals visited, residential socioeconomic deprivation (French Deprivation Index, FDep20; higher values indicate greater deprivation), and predominant ward type. Predominant ward type was defined as the ward type in which each patient accumulated the greatest total length of stay; ward-type labels correspond to PMSI codes. **(B)** Selected hospital-level characteristics: mean length of stay, annual admissions, observed hospital capacity, proportion of patients with more than one admission, number of medical units, and inter-hospital connectivity. Observed hospital capacity was defined as the 95th percentile of daily inpatient census in 2023, and inter-hospital connectivity as hospital degree in the patient-sharing network.

At the hospital level (Fig. 4B), the strongest positive gradient was observed for mean length of stay, with modeled risk increasing across categories and reaching approximately 2.8 times the overall hospital average in the longest-stay category. Higher observed hospital capacity and a greater proportion of patients with more than one admission were also associated with progressively higher risk, reaching approximately 2.0 and 1.9 times the model average, respectively. Annual admissions showed a more moderate increase, with greater uncertainty in the highest-volume category. The number of medical units showed the opposite pattern: after adjustment for the other hospital characteristics, modeled risk decreased progressively as the number of medical units increased. Inter-hospital connectivity showed comparatively modest differences, with most categories remaining close to the model average and moderate elevations among the most highly connected hospitals. Taken together, these results indicate that hospital-level modeled risk was more strongly differentiated by measures of cumulative inpatient exposure, capacity, repeated healthcare use, and internal organization than by inter-hospital connectivity alone. Complete outputs for both models—including the number of patients or hospitals in each category, the corresponding raw model coefficients with reference levels, and the complete set of standardized risk ratios with 95% confidence intervals—are reported in the SI Appendix (Tables S5–S6).

### Sensitivity analyses

The relative structure of simulated colonization risk was robust to uncertainty in colonization duration and initial seeding. Patient-level risk rankings remained highly concordant with the primary analysis when the assumed mean colonization duration was changed from 180 days to 90 or 365 days (Spearman’s *ρ* = 0.987 and 0.988, respectively), and when the number of initially colonized patients was changed from *N*_0_ = 10 to *N*_0_ = 1 or *N*_0_ = 20 (Spearman’s *ρ* = 0.973 and 0.990, respectively; *SI Appendix*, Figs. S6–S5). These assumptions had a greater effect on the overall simulated colonization burden than on the relative ordering of patient-level risk or the broad spatial pattern of risk. Together, these analyses indicated that absolute simulated colonization burden depended on assumptions about colonization duration and initial seeding, whereas the relative pattern of risk across patients remained comparatively stable.

## Discussion

In this study, we developed a nationwide patient-based framework to characterize how simulated hospital-associated colonization risk was distributed across the French acute-care system in relation to observed patient mobility and ward co-location patterns. We reconstructed a temporally resolved network of overnight ward co-location and simulated colonization using a susceptible–colonized–susceptible model allowing clearance and recolonization, with relative transmission multipliers across major clinical groups calibrated using national CPE surveillance data. Simulated risk was highly heterogeneous across patients, ward types, hospitals, and patients’ residential areas, with higher patient-level risk associated particularly with more frequent admissions, longer stays within wards, and visits to more wards. These relative risk patterns remained highly stable when assumptions about colonization duration and initial seeding were varied. At the clinical-group level, simulated and surveillance-derived profiles were strongly rank-concordant, representing post-calibration consistency with the surveillance data used for calibration. At the hospital level, which was not targeted during calibration, higher simulated relative colonization rates were associated with higher odds of recording at least one CPE case in 2023 after adjustment for hospital activity. The simulations characterize the distribution of hospital-associated colonization risk over 365 days following stochastic introduction into the observed hospital network and are not intended to represent the long-term endemic equilibrium or the absolute burden of CPE in France. Overall, these findings show how nationwide patient trajectories can reveal a highly structured distribution of hospital-associated colonization risk across patients and healthcare settings.

A key finding of our study is the contrast between hospital-level and residential-level spatial structure. In our framework, all modeled acquisition occurs during acute-care hospitalization through ward co-location; community and post-discharge transmission are not explicitly simulated. Nevertheless, when simulated hospital-associated colonization risk was aggregated according to patients’ places of residence, clear geographic hotspots emerged, whereas hospital-level risk showed much weaker spatial structure. These residential hotspots should therefore not be interpreted as evidence of local community transmission, but rather as areas whose residents experienced greater colonization risk through their hospital trajectories. The weak association with population density, together with stronger descriptive associations with measures of healthcare use, further suggests that these patterns reflect how populations from different areas interact with the hospital system. Patients living in the same area may use overlapping hospitals and clinical services, allowing hospital-associated risk to become geographically concentrated when mapped back to their places of residence. Importantly, discharge does not necessarily mark the end of potential transmission pathways: colonized patients may subsequently enter rehabilitation centers, long-term care facilities, dialysis services, or other healthcare and community settings where onward transmission could occur [53–55]. Although these settings were not represented in the present network, they provide plausible links between acute-care exposure and the broader continuum of care. From a public health perspective, these findings suggest that considering both patients’ places of residence and their hospital-use patterns may provide additional context for interpreting hospital-associated colonization risk. Such information could help identify patient pathways and groups of healthcare facilities that warrant closer surveillance or further investigation. However, these patterns should not be interpreted as direct intervention targets without pathogen-specific evaluation and consideration of transmission occurring outside the acute-care hospital network.

The multivariable analyses extend the spatial and descriptive findings by showing that simulated colonization risk was associated with characteristics at both the patient and hospital levels. At the patient level, the strongest gradients were related to the frequency and duration of hospital exposure and to predominant ward type, emphasizing differences across patients’ healthcare trajectories and care settings. At the hospital level, measures of cumulative inpatient exposure, capacity, and repeated healthcare use were more strongly associated with modeled risk than inter-hospital connectivity alone, while internal organization showed additional adjusted associations. Together, these findings indicate that simulated colonization risk varies with both patients’ trajectories through the hospital system and characteristics of the facilities in which care occurs. From a public health perspective, surveillance and infection-prevention efforts may therefore benefit from considering high-exposure patient trajectories and hospital settings with greater cumulative patient exposure, rather than assuming comparable risk across patients and facilities.

The hospital-level comparison with CPE surveillance provided an additional empirical consistency check because individual hospitals were not targeted during calibration. Higher simulated relative colonization rates were associated with a greater likelihood of recorded CPE occurrence, even after accounting for differences in hospital activity. This suggests that the simulated hospital-level risk measure contained information associated with recorded CPE occurrence beyond hospital activity alone. However, the comparison was based only on whether a hospital recorded at least one CPE case and should not be interpreted as validation of hospital-specific CPE incidence or case burden.

Several limitations should be considered. First, although clinical-group-specific transmission probabilities captured broad heterogeneity across care settings, the model did not represent differences between individual hospitals or patients arising from antimicrobial exposure, infection-prevention practices, staffing, case mix, or individual susceptibility. The simulated probabilities should therefore be interpreted as relative measures of hospital-associated colonization risk rather than estimates of absolute CPE prevalence. Second, the network was restricted to acute-care inpatient pathways. Long-term care facilities, rehabilitation centers, nursing homes, dialysis services, and other outpatient settings were not represented, potentially omitting important transmission pathways and connections between acute-care hospitals and the broader continuum of care [53–56]. Third, ward-level overnight co-location represents a plausible transmission opportunity rather than direct physical contact. PMSI does not provide room- or bed-level proximity, healthcare-worker contacts, or environmental exposures; the ward-based definition may therefore overestimate relevant contact in some settings while missing shorter exposures that may be important for particular pathogens. We did not test alternative contact definitions; pathogen-specific applications may require broader or time-lagged exposure definitions. In addition, acquisition and transmission outside hospitals were not simulated, so residential risk patterns should not be interpreted as estimates of community transmission. Finally, e-SIN surveillance is not exhaustive: screening, laboratory detection, and reporting practices vary across hospitals, and the absence of a recorded case does not imply the absence of CPE [57].

Despite these limitations, our study provides several clear directions for further work. First, the network could be extended beyond acute-care hospitals to include long-term care, rehabilitation, dialysis, and other healthcare settings, allowing patient pathways and potential transmission to be followed across a broader continuum of care. Second, incorporating hospital-specific modifiers, such as antimicrobial use or infection-prevention and control practices, could allow transmission risk to vary between hospitals in addition to the clinical-group differences represented in the present model. Third, the PBN could be used to test alternative introduction scenarios, including seeding at different times or seasons and in specific hospitals, clinical settings, or geographic areas. Finally, comparing the patient-based network with coarser ward- and hospital-level networks derived from the same data could show which risk patterns are preserved across levels of aggregation and when individual-level detail is necessary. Comparable national hospitalization databases in other countries, such as Hospital Episode Statistics in the United Kingdom [58], could also support adaptation of this framework to other healthcare systems. Overall, this framework provides a scalable approach for linking patient-mobility networks with pathogen-specific surveillance to identify where hospital-associated colonization risk concentrates across healthcare systems.

## Materials and Methods

### Data Description

This study combined two complementary national data sources: nationwide hospital-discharge records used to reconstruct patient trajectories and national surveillance data used to inform a pathogen-specific application for carbapenemase-producing Enterobacterales (CPE). Patient trajectories were reconstructed from the 2023–2024 PMSI–MCO database (*Programme de Médicalisation des Systèmes d’Information–Médecine, Chirurgie, Obstétrique*), France’s national hospital-discharge database administered by the Technical Agency for Information on Hospitalization (ATIH). PMSI–MCO records inpatient and outpatient activity in public and private acute-care hospitals, including admission and discharge dates, hospital and medical-unit identifiers, patient characteristics, diagnoses, and ward-level movements. Stable pseudonymized identifiers allowed successive stays and movements between wards to be linked over time. In 2023–2024, the database contained approximately 63 million hospital stays for more than 22.6 million distinct patients across more than 1,500 acute-care hospital sites. We restricted the analysis to the acute-care medicine–surgery–obstetrics sector and excluded the psychiatry (PMSI– PSY), rehabilitation (*Soins de Suite et de Réadaptation*; PMSI–SSR), and home-hospitalization (*Hospitalisation à Domicile*; PMSI–HAD) sectors. Comparable day-resolved medical-unit trajectories were not available to us across these non-acute-care sectors, preventing consistent application of the same ward co-location definition used for the acute-care PBN. Hospital sites in overseas departments were also excluded, so the study covered metropolitan France only (96 administrative departments). Detailed source files, linkage procedures, and eligibility criteria are described in the *SI Appendix* and Fig. S1.

CPE surveillance data were obtained from e-SIN, the French electronic reporting system for healthcare-associated infection and antimicrobial-resistance events. One e-SIN *signalement* represents a surveillance episode and may include one or several colonized or infected cases and one or more associated hospital services. We used national reports of highly resistant emerging bacteria (*bactéries hautement résistantes émergentes*, BHRe) recorded from 2017 through 2025 and retained CPE episodes reported by metropolitan acute-care hospitals. Hospital services were mapped to the 12 clinical groups used in the PMSI analysis. After applying the surveillance eligibility criteria, the final dataset comprised 12,350 CPE episodes and 18,883 reported cases. Pooled 2017–2025 data were used to inform clinical-group transmission heterogeneity, whereas hospital-level reports from 2023 were used separately to assess consistency between simulated risk and recorded CPE occurrence. Detailed surveillance processing is provided in the *SI Appendix*, Section S1.1 and Fig. S2.

### Patient-based network construction

To construct the patient-based network (PBN), we extracted all acute-care stays recorded in PMSI–MCO between January 1, 2023, and December 31, 2024, together with early-2025 discharge records for patients admitted in late 2024. Because PMSI is discharge based, stays ending in 2025 appear only in the 2025 extract. Including these records ensured complete hospitalization trajectories for all admissions beginning during 2023–2024. We applied sequential eligibility criteria to retain plausible and consistently measurable opportunities for hospital-based transmission (Fig. S1). Records with invalid or placeholder patient identifiers were excluded because they could not be reliably linked across stays. Outpatient and day-hospital activity, including dialysis, radiotherapy, and chemotherapy, was excluded because patients are generally present for only part of the day and PMSI does not provide sufficiently precise within-day timing to establish simultaneous presence in the same medical unit. Some dialysis records also contained implausibly long durations, indicating potential data-quality problems. We therefore defined a potential contact as co-location in the same medical unit for at least one overlapping night. This provided a consistent measure of sustained ward occupancy and a plausible opportunity for direct proximity or indirect exposure through healthcare personnel, shared equipment, or the ward environment. This definition does not imply that transmission cannot occur during shorter encounters, but restricts the network to contacts that can be identified reliably and comparably across hospitals. From the remaining trajectories, we identified all pairs of patients who shared at least one overlapping night in the same medical unit (ward). We constructed an undirected temporal PBN in which nodes represented individual patients and time-stamped edges represented overnight ward co-location. The final 2023–2024 PBN comprised 10,413,176 patients, 326,639,314 pairwise ward-level co-location interactions, and 1,408 hospitals in metropolitan France. A detailed flowchart of the data-cleaning procedures, the construction of PBN-France, and the heterogeneity observed in the static network analyses (e.g., degree distribution) is provided in the *SI Appendix*, Fig. S1 and S3.

### Stochastic SCS transmission model and simulation design

To evaluate how healthcare-associated CPE colonization may propagate through the French hospital system, we simulated transmission on the nationwide temporal PBN described above using a stochastic susceptible–colonized–susceptible (SCS) model. For each calendar day *t*, the corresponding network layer contained all overnight ward co-location contacts observed on that day. Because these layers changed with new admissions, discharges, ward transfers, and movements between hospitals, the simulations preserved the observed temporal organization of patient flows across the hospital system. Each patient was classified as susceptible (*S*) or colonized (*C*). Colonized patients could transmit to susceptible ward contacts, subsequently clear colonization and return to the susceptible state, and later become recolonized.

Every ward retained in the PBN was assigned to one of 12 clinical groups. Transmission remained defined between patients co-located in the same ward, but the daily per-neighbor transmission probability depended on the clinical group of that ward:

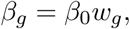

where *g* indexes the 12 clinical groups, *β*_0_ = 0.05 is the baseline transmission probability used as a reference scale for parameterizing clinical-group differences, and *w_g_* is the calibrated multiplier for clinical group *g*. The baseline value *β*_0_ was not interpreted as a direct biological estimate of CPE transmissibility; the resulting clinical-group-specific transmission probability was determined by *β_g_* = *β*_0_*w_g_*, as described in the next subsection. Clinical-group heterogeneity therefore modified transmission probabilities without changing the underlying ward-level contact network. Let *k_j_*(*t*) denote the number of colonized patients sharing the ward of susceptible patient *j* on day *t*, and let *g*(*j, t*) denote the clinical group of that ward. Assuming independent transmission opportunities across colonized neighbors, the probability that patient *j* acquired colonization on day *t* was

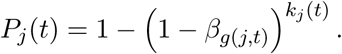

For each susceptible patient, we generated *U_j_*(*t*) ∼ Uniform(0, 1), and colonization occurred when *U_j_*(*t*) *< P_j_*(*t*). State transitions were implemented synchronously: transmission and clearance outcomes on day *t* were determined from patient states at the beginning of that day and applied before the next daily network layer. Consequently, only patients colonized at the beginning of day *t* could transmit on that day, whereas patients newly colonized on day *t* could begin transmitting on day *t* + 1.

Colonized patients cleared colonization independently each day with probability *γ* = 1 – 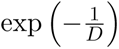, where *D* was the assumed mean colonization duration. The primary analysis used *D* = 180 days, corresponding to *γ* ≃ 0.00554 per day. This value was selected as a plausible intermediate assumption within the range of CPE carriage durations reported in previous studies [6, 59]. Sensitivity analyses considered *D* = 90 and *D* = 365 days to represent shorter and longer persistence of colonization. Clearance continued throughout calendar time, including periods outside hospital. Patients outside hospital had no PBN contacts and could therefore neither acquire nor transmit colonization through the hospital network, but they could clear colonization before a subsequent admission. Patients readmitted before clearance remained colonized, whereas those who cleared returned to the susceptible state and could subsequently become recolonized.

At the start of each simulation, 10 patients were randomly selected from those present in the network on the sampled start date and assigned to the colonized state; all other patients were initially susceptible. Ten was chosen as a standardized national seeding level comparable in order of magnitude to the median number of BHRe cases reported per day through e-SIN in 2023. This value was not interpreted as an estimate of prevalent CPE carriage. Sensitivity analyses using 1 and 20 initially colonized patients assessed the influence of this assumption. Simulation start dates were randomly sampled from 2023, and each run was followed for 365 consecutive days using the observed sequence of daily network layers across 2023–2024. After calibration, the clinical-group multipliers were fixed, and the primary model was repeated 2,000 times using independently sampled start dates and initial colonized patients.

### Surveillance-informed calibration of clinical-group transmission

For calibration, we used the processed e-SIN surveillance cohort to estimate the clinical-group transmission multipliers *w_g_* so that the simulated relative distribution of CPE burden across the 12 clinical groups reflected the surveillance-derived profile. When an episode included several reported cases and several clinical groups, its cases were divided equally among the unique groups represented, preventing double counting while preserving the total reported-case count. For each clinical group *g*, the calibration target was defined as its share of weighted reported CPE cases divided by its share of national PMSI patient-days in 2023. Patient-days were defined as the total number of days spent by all patients in each clinical group and therefore accounted for differences in inpatient exposure between groups. A target value above one indicated that a clinical group contributed a larger share of reported CPE cases than expected from its share of inpatient exposure, whereas a value below one indicated lower representation. The 2023 PMSI patient-day distribution was used because the primary simulated cohort comprised patients hospitalized in 2023; the corresponding clinical-group distribution was nearly identical in 2024. Construction of the surveillance cohort and calibration targets is presented in the *SI Appendix* and Fig. S2.

We calibrated the 12 clinical-group transmission multipliers (*w_g_*) so that the relative distribution of simulated colonizations across clinical groups resembled that observed in CPE surveillance. For each candidate multiplier vector, **w** = (*w*_1_*,…, w*_12_), we ran the same 10 predefined 365-day simulations and counted first model-generated, non-seed colonizations occurring in each clinical group. Both the simulated and surveillance profiles were expressed relative to each group’s share of national PMSI patient-days in 2023, allowing them to be compared on the same scale. The derivative-free Subplex algorithm, implemented in the R package nloptr [60], searched the bounded parameter space by evaluating successive candidate vectors and retained the best-performing vector identified according to a penalized loss that balanced agreement between the two 12-group profiles against extreme multiplier values. Calibration was performed separately for mean colonization durations of 90, 180, and 365 days. The resulting multipliers represent surveillance-informed differences between clinical settings and should not be interpreted as direct biological estimates of CPE transmissibility. Full calibration details are provided in the *SI Appendix*, Section S1.2, and the clinical-group-specific values used in the primary 180-day analysis are reported in Table S4.

### Colonization-risk aggregation from simulation outcomes

From each of the 2,000 primary SCS simulation runs, we recorded for every patient (i) whether they experienced a first model-generated, non-seed colonization, (ii) the day of the 365-day follow-up on which it occurred, and (iii) the hospital and ward type in which the event occurred. Although clearance and recolonization were included in the SCS dynamics, only the first non-seed colonization was retained as the patient-level outcome. Thus, within a given run, each patient was assigned at most one hospital–ward-type acquisition location. Across runs, however, the same patient could acquire colonization in different locations because the stochastic transmission process could unfold differently. Patient-level probabilities and all subsequent risk analyses were restricted to patients hospitalized at least once in 2023, because simulation start dates were sampled from 2023 and the two-year network provided complete 365-day follow-up for these patients.

#### Patient-level colonization probabilities

For patient *j*, let *n_jℓ_* denote the number of eligible simulations in which the patient experienced a first non-seed colonization at hospital–ward-type location *ℓ*. Let *M_j_*denote the number of simulations in which patient *j* was not selected as an initial colonized patient. The location-specific colonization probability was

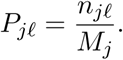

Thus, simulations in which patient *j* was selected as an initial seed were excluded from that patient’s denominator because they provided no opportunity to observe a model-generated first acquisition.

From the location-specific probabilities *P_jℓ_*, we derived the patient-level quantities used in subsequent analyses. The overall patient colonization probability, *P_j_* = Σ*_ℓ_ P_jℓ_,* represented the probability that patient *j* experienced a first model-generated, non-seed colonization during follow-up, regardless of acquisition location. Because only the first colonization was retained within each simulation, location-specific acquisition events were mutually exclusive within a run and could therefore be summed. The hospital-attributed colonization probability was *P_j__H_* = Σ *_ℓ∈H_ P_jℓ_,* and the ward-type-attributed probability *P_jk_* was obtained by summing *P_jℓ_* across hospitals for locations belonging to ward type *k*. Clinical-group-attributed probabilities were obtained analogously by summing across ward types belonging to the same clinical group. A worked numerical example is provided in the *SI Appendix*, Section S1.3. These patient-level probabilities were then averaged at the corresponding aggregation level. Residential-area risk was the mean of *P_j_* among patients residing in the same area; hospital-level risk was the mean of *P_j__H_* among patients who visited hospital H; and ward-type- and clinical-group-level risks were the means of their corresponding attributed probabilities among patients who visited those settings.

#### Colonization risk ratios

For comparisons across residential areas, hospitals, ward types, and clinical groups, each mean colonization probability was expressed as a ratio to the corresponding national mean at the same aggregation level. A risk ratio of 1 therefore represents the national average, with values above or below 1 indicating higher or lower simulated colonization risk, respectively.

### Spatial and statistical analysis

#### Spatial autocorrelation analysis

To assess whether simulated colonization risk exhibited non-random spatial structure, we quantified spatial autocorrelation at two levels: patients’ residential areas and hospital locations. For residential analyses, we used the PMSI residential geographic code, an ATIH-defined, postal-code-based geographic identifier derived from patients’ place of residence; some postal codes are grouped according to ATIH geographic coding rules. Patients sharing the same residential geographic code were assigned to the same residential unit. After restricting to valid residential locations in metropolitan France, 5,533 unique residential units remained. One Voronoi cell was constructed from the geographic location associated with each residential unit and assigned the corresponding residential colonization risk ratio. For hospital analyses, we first grouped geographically proximate hospitals using DBSCAN with a 10 km radius to improve map readability. For each hospital cluster, colonization risk was summarized as the weighted mean of the hospital-level risks, using the number of PBN patients associated with each hospital as weights. We then constructed Voronoi polygons over the resulting hospital clusters and assigned each polygon the corresponding cluster-level colonization risk ratio [61]. At both levels, overall spatial dependence was quantified using Global Moran’s *I*, and localized clustering was assessed using Local Indicators of Spatial Association (LISA), which identified high–high hotspots, low–low coldspots, and spatial outliers. In both analyses, spatial weights were based on queen contiguity (polygons sharing an edge or a vertex) and row-standardized before analysis. Statistical significance for Global Moran’s *I* and LISA was assessed using standard spatial-statistical tests; full details are provided in the *SI Appendix*, Section S1.4.

#### Multivariable models of colonization risk

To identify patient- and hospital-level characteristics associated with simulated colonization risk, we fitted separate multivariable beta-regression models with a logit link using mgcv::bam [62]. Models were fitted separately at the patient and hospital levels using REML-based estimation. At the patient level, the outcome was each patient’s overall simulated colonization probability, *P_j_*. Covariates were organized into five complementary dimensions: patient characteristics (age and sex); healthcare use (number of admissions and mean length of stay per ward); movement through the hospital system (numbers of hospitals and wards visited and mean residence-to-hospital distance); predominant care environment (ward type in which the patient accumulated the greatest total length of stay); and residential context (population density and socioeconomic deprivation). Mean residence-to-hospital distance was calculated across hospitals visited, weighted by the number of admissions to each hospital. Residential socioeconomic context was characterized using the French Deprivation Index (FDep20), an area-level index calculated as the negative first principal component of four 2020 municipality-level indicators—median income per consumption unit, educational attainment, proportion of manual workers, and unemployment—with higher values indicating greater socioeconomic deprivation [63]. The patient-level model was fitted using complete cases without imputation (*n* = 5,849,046 of 5,910,857 eligible patients). At the hospital level, the outcome was hospital-level simulated colonization risk, defined as the mean of the patient-specific hospital-attributed colonization probabilities, *P_j__H_*, across patients who visited hospital H. Covariates represented four complementary dimensions of hospital organization and patient flow: activity and capacity (annual admissions, observed hospital capacity, and number of medical units); care organization (mean length of stay and number of ward types); repeated healthcare use (proportion of patients with more than one admission); and position in the national patient-sharing network (inter-hospital connectivity). Observed hospital capacity was defined as the 95th percentile of daily inpatient census in 2023, and inter-hospital connectivity as hospital degree in the patient-sharing network. Several strongly skewed continuous predictors were grouped into ordered categories to retain interpretable ranges while limiting sparsely populated groups. Exact category definitions and beta-regression boundary-value handling are provided in the *SI Appendix*, Section S1.6.

To present model output on an interpretable risk scale, we estimated adjusted predicted risks for each predictor category by marginal standardization and expressed them as standardized risk ratios (SRRs) [64]. For each model separately, the SRR was calculated as the category-specific adjusted predicted risk divided by the overall mean predicted risk from the same model, such that SRR = 1 represents the model-specific average risk. SRRs were therefore used for within-model comparisons across patient- and hospital-level characteristics. Full standardization procedures and complete model outputs are provided in the *SI Appendix*, Section S1.6 and Tables S5–S6.

## Data availability

The datasets analyzed in this study are owned by third parties and are not publicly available due to hospital and patient privacy. ATIH owns the PMSI–MCO database. Researchers may request access directly from ATIH (process described at www.atih.sante.fr). Santé publique France (SpF) owns the CPE surveillance data from e-SIN. Researchers may request access directly from SpF (process described at www.santepubliquefrance.fr). From these routinely collected pseudonymized hospital-discharge records, we reconstructed a fully anonymized patient-based network. All transmission analyzes were simulation-based. The study did not involve prospective patient recruitment, direct patient contact, or study-specific intervention.

## Acknowledgments

We thank the ARCANE project members for valuable discussions, ATIH for providing access to PMSI data, and Santé publique France for providing access to e-SIN CPE surveillance data. We acknowledge the GenOuest bioinformatics core facility (https://www.genouest.org) for providing high-performance computing resources. We also thank the Editor and the three anonymous reviewers for their thoughtful and constructive comments, which helped strengthen the manuscript.

## SI Appendix

### S1. Supplementary Methods

### S1.1 Processing of e-SIN surveillance data

We used national e-SIN (External Reporting of Healthcare-Associated Infections) reports of highly resistant emerging bacteria (*bactéries hautement résistantes émergentes*, BHRe) recorded from 2017 through 2025. One *signalement* represented one surveillance episode and could include one or several colonized or infected cases and one or more associated hospital services. The initial dataset contained 17,990 BHRe episodes and 28,075 reported cases. Reports were sequentially restricted to hospital sites in metropolitan France, acute-care/MCO hospitals, and episodes with at least one reported service that could be mapped to one of the 12 clinical groups used in the patient-based network. An episode was classified as carbapenemase-producing Enterobacterales (CPE) only when an Enterobacterales organism and a specific carbapenemase mechanism were reported for the same microorganism entry. The retained carbapenemase families included KPC, NDM, VIM, IMP, IMI, and OXA-48-like enzymes. Episodes associated with direct medical repatriation from abroad were excluded because they represented likely external importations rather than acquisitions occurring within the French hospital network. After these restrictions, the final analytical cohort comprised 12,350 CPE episodes and 18,883 reported cases. The sequential filtering steps and numbers retained at each stage are shown in Fig. S2A.

#### S1.2 Calibration of clinical-group transmission multipliers

##### Construction of the surveillance targets

Hospital services reported in e-SIN were mapped to the same 12 clinical groups used to classify PMSI wards. Because one surveillance episode could include several reported cases and several hospital services, duplicate occurrences of the same clinical group within an episode were first removed. For an episode *e* containing *C_e_* reported cases and *m_e_* unique clinical groups, each represented group was allocated *C_e_/m_e_* cases. These allocations summed to *C_e_* within each episode, thereby preventing double counting while preserving the total reported-case count. For the episode-based robustness comparison, each episode was also assigned a total weight of one, divided equally among its *m_e_* unique clinical groups; each represented group therefore received a weight of 1*/m_e_*. The reported-case-based profile was used for calibration because the simulated outcomes were individual first colonization events, whereas the episode-based profile was used only to assess robustness to the surveillance quantity selected.

For each clinical group *g*, the weighted reported-case count was converted into its share of all reported CPE cases. To account for differences in inpatient exposure between clinical groups, this share was divided by the corresponding share of national PMSI patient-days in 2023. One patient-day corresponded to one patient spending one day in a given clinical group, and group-specific patient-days were obtained by summing these days across all patients in 2023. The surveillance target for clinical group *g* was therefore defined as

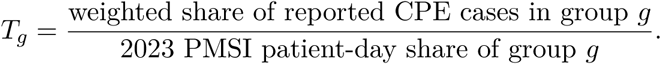

A value of *T_g_ >* 1 indicated that the clinical group accounted for a larger share of reported CPE cases than expected from its share of inpatient exposure, whereas *T_g_ <* 1 indicated lower representation relative to inpatient exposure. The same 2023 PMSI patient-day shares were subsequently used to standardize the simulated distribution of first colonizations, ensuring that the surveillance and simulated ratios were directly comparable. The 2023 denominator was selected because simulations were initiated in 2023 and the calibration cohort comprised patients hospitalized during that year. Clinical-group patient-day shares were nearly identical in 2023 and 2024 (Pearson *r* = 0.9996; Spearman *ρ* = 1.00). The episode- and reported-case-based standardized profiles were also strongly concordant (Pearson *r* = 0.98; Spearman *ρ* = 0.97).

##### Transmission parameters and calibration scenarios

For clinical group *g*, the daily per-neighbor transmission probability was *β_g_* = *β*_0_*w_g_,* where the baseline probability was fixed at *β*_0_ = 0.05 and *w_g_* was the clinical-group-specific multiplier. The 12 positive multipliers were optimized on the logarithmic scale: *θ_g_*= log(*w_g_*)(*or w_g_* = exp(*θ_g_*)). This transformation ensured that all proposed multipliers remained positive. During optimization, the multipliers were constrained to 0.10 ≤ *w_g_* ≤ 4.50. This broad range allowed the optimizer to represent the substantial relative heterogeneity observed in the surveillance-standardized clinical-group profiles (Fig. S2C), while preventing exploration of excessively small or large transmission parameters. Given *β*_0_ = 0.05, the constraints corresponded to daily per-neighbor transmission probabilities between 0.005 and 0.225. For the primary 180-day analysis, the final calibrated multipliers ranged from *w_g_* = 0.100 to 4.467, corresponding to daily per-neighbor transmission probabilities of *β_g_* = 0.0050 to 0.2233; group-specific values are reported in Table S4.

Each candidate multiplier vector, **w** = (*w*_1_*,…, w*_12_), was evaluated using *n*_cal_ = 10 fixed 365-day calibration simulations. These scenarios were generated once before optimization, with each scenario having a prespecified start date, a fixed set of 10 initially colonized patients, and a scenario-specific random-number seed. The same scenarios and random-number seeds were used for every candidate vector, reducing stochastic variation between candidate evaluations so that differences in the objective more directly reflected changes in the proposed multipliers. The use of 10 scenarios was a pragmatic compromise between reducing simulation variability and computational cost. These fixed scenarios were used only during calibration; the final simulation ensemble used independently sampled start dates and initially colonized patients.

##### Simulated clinical-group profile

For each calibration simulation, we counted the first model-generated, non-seed colonizations occurring in each clinical group. Initially colonized patients were excluded because their colonization was assigned at the start of the simulation, and subsequent recolonizations were not counted as additional calibration outcomes. Let *A_gs_* denote the number of first non-seed colonizations occurring in clinical group *g* in calibration simulation *s*. Within each simulation, these counts were converted to clinical-group shares as

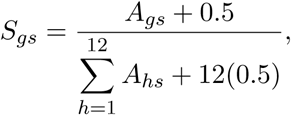

where the pseudocount of 0.5 prevented zero-valued shares for clinical groups with no acquisitions in a given simulation. The simulation-specific patient-day-standardized ratio was then

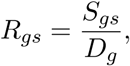

where *D_g_*denotes clinical group *g*’s share of national PMSI patient-days in 2023, using the same exposure denominator as in the surveillance target.

A value of *R_gs_ >* 1 indicated that clinical group *g* accounted for a larger share of simulated first colonizations than expected from its share of inpatient exposure in simulation *s*, whereas a value below 1 indicated lower representation. The ratios were then averaged across the 10 fixed calibration simulations,

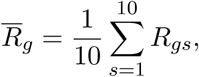

and *R̄_g_* was compared with the corresponding surveillance target *T_g_*.

##### Calibration objective

For each clinical group *g*, we compared the mean simulated acquisition-to-exposure ratio *R̄_g_* with the corresponding surveillance target *T_g_*. Their discrepancy was calculated on the logarithmic scale:

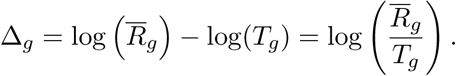

A value of Δ*_g_* = 0 indicated exact agreement. A positive value indicated that the simulation produced a higher ratio than observed in surveillance, whereas a negative value indicated a lower ratio. The logarithmic scale treated proportional deviations symmetrically; for example, a simulated ratio twice the target and a ratio one-half of the target produced discrepancies of equal magnitude but opposite sign.

The optimizer minimized the following penalized objective:

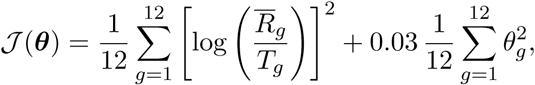

where *θ_g_* = log(*w_g_*). The first term measured the overall mismatch between the simulated and surveillance-derived profiles across the 12 clinical groups. Squaring the discrepancies ensured that deviations above and below the targets both increased the loss, with larger discrepancies receiving greater penalty. All clinical groups contributed equally. The second term discouraged unnecessarily extreme multipliers by favouring values closer to the reference value *w_g_* = 1. Lower values of J therefore indicated a better balance between agreement with the surveillance targets and avoidance of extreme parameter values.

##### Optimization, stopping criteria, and duration-specific calibration

Because the calibration objective could only be evaluated by running the stochastic transmission simulations and analytical derivatives with respect to the 12 multipliers were not available, we used a derivative-free optimization method. Specifically, the penalized objective was minimized using the bounded Subplex algorithm implemented in the R package nloptr through the NLopt routine NLOPT_LN_SBPLX [60]. Subplex performs a local search by evaluating the objective at successive candidate parameter vectors without requiring gradient information. At each evaluation, the proposed 12-dimensional multiplier vector was assessed using the same 10 fixed 365-day calibration simulations described above. Optimization was initialized from the equal-multiplier vector (*w_g_* = 1 for all 12 clinical groups). Optimization stopped when the maximum number of evaluations, an absolute tolerance of 0.01 in the log-multipliers, or the allocated computation-time limit was reached. The multiplier vector with the lowest penalized objective value encountered during the search was retained, and continuation stages were restarted from this vector when necessary. Approximately 300 candidate vectors were evaluated for each colonization duration, corresponding to approximately 3,000 calibration simulations per duration. For the primary 180-day scenario, the penalized objective decreased from 1.1465 for the initial equal-multiplier vector to 0.1176 for the lowest-loss vector retained, corresponding to an approximately 89.7% reduction. Calibration was performed separately for mean colonization durations of *D* = 90, 180, and 365 days because changing the clearance rate altered the simulated distribution of colonization events. The resulting multipliers represent effective, surveillance-informed differences among clinical settings conditional on *β*_0_ = 0.05, rather than direct biological estimates of CPE transmissibility.

#### S1.3 Worked Example of Simulation Output Aggregation

This section provides a simple numerical example illustrating how raw simulation outputs were aggregated into patient-level colonization probabilities, using one patient, two hospitals, four hospital–ward-type locations, and seven simulation runs. All values are hypothetical and chosen for clarity.

##### Setup

Patient *A* visited two hospitals during the year: hospital *H*_1_ (ward types *W*_1_ and *W*_2_) and hospital *H*_2_ (ward types *W*_1_ and *W*_3_). Ward type *W*_1_ therefore occurred in both hospitals, which is relevant for the ward-type aggregation in Step 5.

##### Step 1 — Raw simulation output

For each simulation, only the first model-generated, non-seed colonization of Patient *A* was retained for patient-level risk estimation. Although clearance and subsequent recolonization could occur and continued to affect the transmission dynamics, later colonization events within the same run were not counted as additional patient-level outcomes.

Table S1 shows the location of Patient *A*’s first model-generated colonization across the seven runs. Because the stochastic transmission process could unfold differently between runs, the first colonization could occur at different hospital–ward-type locations.

**Table S1:** Raw simulation output for Patient *A* across seven runs. “—” indicates that no model-generated colonization occurred in that run.

| Run | Colonized? | Location of first colonization |
| --- | --- | --- |
| 1 | Yes | $\mathcal{H}_1, W_1$ |
| 2 | Yes | $\mathcal{H}_1, W_2$ |
| 3 | Yes | $\mathcal{H}_1, W_1$ |
| 4 | No | — |
| 5 | Yes | $\mathcal{H}_2, W_3$ |
| 6 | Yes | $\mathcal{H}_2, W_1$ |
| 7 | No | — |

##### Step 2 — Location-specific colonization probability

*P_jℓ_* For each hospital–ward-type acquisition location *ℓ*, let *n_jℓ_* denote the number of eligible simulation runs in which the first model-generated colonization of patient *j* was attributed to that location. The location-specific colonization probability was

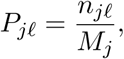

where *M_j_* is the number of simulation runs in which patient *j* was not selected as an initial colonized patient. In this hypothetical example, Patient *A* was not an initial seed in any of the seven runs, so *M_A_* = 7. The resulting location-specific probabilities are shown in Table S2.

**Table S2:** Location-specific colonization probabilities for Patient *A*. Each row corresponds to a distinct hospital–ward-type acquisition location in which the patient’s first model-generated colonization occurred in at least one simulation run.

| Ward type | Hospital | $n_{A\ell}$ | $P_{A\ell} = n_{A\ell}/7$ |
| --- | --- | --- | --- |
| $W_1$ | $\mathcal{H}_1$ | 2 | $2/7 = 0.286$ |
| $W_2$ | $\mathcal{H}_1$ | 1 | $1/7 = 0.143$ |
| $W_3$ | $\mathcal{H}_2$ | 1 | $1/7 = 0.143$ |
| $W_1$ | $\mathcal{H}_2$ | 1 | $1/7 = 0.143$ |

Patient *A* therefore contributed four distinct patient–hospital–ward-type acquisition records. More generally, a patient could contribute to several locations across simulations when the location of first colonization differed between runs.

##### Step 3 — Overall patient colonization probability

*P_j_* Because only one first model-generated colonization was retained for a patient within each simulation, the location-specific acquisition events were mutually exclusive within a run and could therefore be summed to obtain the overall patient colonization probability:

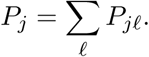

For Patient *A*,

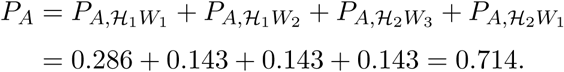

This agrees directly with Table S1, in which Patient *A* experienced a model-generated colonization in five of seven eligible runs (5*/*7 = 0.714).

##### Step 4 — Hospital-attributed colonization probability

*P_jH_* The patient colonization probability attributed to hospital H was obtained by summing the location-specific probabilities over all acquisition locations within that hospital:

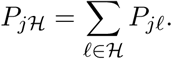

For Patient *A*,

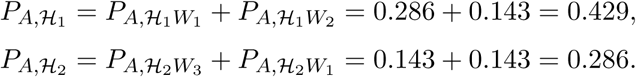

Thus, the same patient could contribute hospital-attributed probabilities to more than one hospital across simulation runs.

##### Step 5 — Ward-type-attributed colonization probability

*P_jk_* The patient colonization probability attributed to ward type *k* was obtained by summing the location-specific probabilities across all hospitals in which the corresponding ward type occurred:

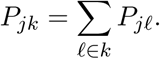

This is relevant when the same ward type occurs in more than one hospital. For Patient *A*, ward type *W*_1_ occurred in both H_1_ and H_2_:

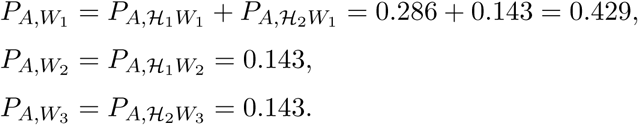

The resulting ward-type-attributed probabilities are summarized in Table S3.

**Table S3:** Ward-type-attributed colonization probabilities for Patient *A*, obtained by summing location-specific probabilities across hospitals for each ward type.

| Ward type | Hospitals represented | $P_{Ak}$ |
| --- | --- | --- |
| $W_1$ | $\mathcal{H}_1, \mathcal{H}_2$ | $0.286 + 0.143 = 0.429$ |
| $W_2$ | $\mathcal{H}_1$ | 0.143 |
| $W_3$ | $\mathcal{H}_2$ | 0.143 |

Together, these steps illustrate how the first model-generated colonization recorded in each simulation was converted into overall, hospital-attributed, and ward-type-attributed patient colonization probabilities used in the subsequent analyses.

#### S1.4 Spatial autocorrelation analysis of colonization risk (Global and Local Moran’s *I*)

Autocorrelation measures how strongly a variable is correlated with itself when observations are separated in time (temporal autocorrelation) or in space (spatial autocorrelation). Spatial autocorrelation describes the extent to which values observed at nearby locations are more similar (or more different) than would be expected by chance. This idea is closely related to Tobler’s First Law of Geography: “Everything is related to everything else, but near things are more related than distant things” [65]. Positive spatial autocorrelation arises when neighbouring regions tend to have similar values (spatial clustering), whereas negative spatial autocorrelation arises when neighbours tend to have opposite values (spatial dispersion).

In this study, we used spatial autocorrelation to assess whether simulation-derived colonization risk exhibited non-random spatial structure across France—i.e., whether risk varied systematically between locations and whether high- and low-risk areas tended to cluster rather than occurring independently in space. We performed the analysis at two spatial scales: (i) residential risk, using Voronoi polygons centered on patients’ residential codes (codeGeo); and (ii) hospital-level risk, using Voronoi polygons constructed over spatial clusters of hospitals identified using DBSCAN (10 km radius; eps=10,000 m, minPts=1) to improve map readability. For each cluster, colonization risk was calculated as the weighted mean of the hospital-level risks, using the number of PBN patients associated with each hospital as weights. Spatial dependence was quantified using Global Moran’s *I* (overall spatial autocorrelation) and local indicators of spatial association (LISA; spatial clustering and outliers).

##### Global Moran’s *I*

To test whether colonization risk is spatially structured rather than randomly distributed, we calculated the Global Moran’s *I* statistic for each risk surface (residential Voronoi polygons and hospital Voronoi polygons), following standard spatial-statistics practice [66, 67]. For an attribute *Y_i_*observed in *n* spatial units, Moran’s *I* is defined as

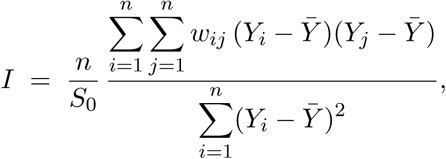

where *Ȳ* is the mean of *Y* across all spatial units, *w_ij_*is the spatial weight between units *i* and *j*, and

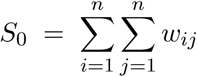

is the sum of all spatial weights. In our setting, *Y_i_*is the colonization risk ratio in polygon *i*, and *w_ij_* = 1 when polygons *i* and *j* share an edge or a vertex (queen contiguity) and *w_ij_* = 0 otherwise; the weights are then row-standardized.

Intuitively, Moran’s *I* compares how similar neighbouring units are to how variable the dataset is overall. If neighbouring polygons tend to have similar values (both above or both below the mean), the cross-products (*Y_i_* − *Y*^̄^)(*Y_j_* − *Y*^̄^) are positive and *I >* 0, indicating positive spatial autocorrelation. If neighbours tend to have opposite values (one high, one low), the cross-products are negative and *I <* 0, indicating negative spatial autocorrelation. Values of *I* ≈ 0 are consistent with no spatial structure. The null hypothesis is spatial independence: colonization risk is randomly arranged in space, given the marginal distribution of *Y*. We computed Moran’s *I*, its expected value, and its variance using moran.test from the spdep package and assessed statistical significance using the corresponding normal approximation (z-score and *p*-value) [68].

##### Local Moran’s *I* (LISA)

Global Moran’s *I* summarizes spatial dependence in a single statistic and does not indicate *where* clusters occur. To identify specific areas that contribute most to the global pattern, we computed Local Indicators of Spatial Association (LISA), i.e., the local Moran statistic *I_i_* for each polygon [67, 69]. For an attribute *Y_i_*observed in *n* spatial units, the local Moran’s *I* at unit *i* is

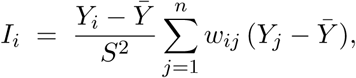

where *Y*^̄^ is the mean of *Y*, *w_ij_* are elements of the spatial-weights matrix, and

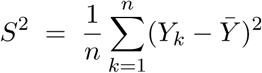

is the variance normalization term for *Y* [70]. As above, *Y_i_* is the colonization risk ratio in polygon *i*, and *w_ij_* = 1 when polygons *i* and *j* share an edge or a vertex (queen contiguity) and *w_ij_* = 0 otherwise; the weights are row-standardized before computing *I_i_*. Large positive values of *I_i_* indicate that unit *i* has a value similar to those of its neighbours (a local cluster), whereas large negative values indicate that unit *i* is an outlier relative to its neighbours (high surrounded by low, or vice versa). Statistical significance for each *I_i_* was assessed using the standard normal-approximation test implemented in localmoran from the spdep package. This procedure returns, for each polygon, the local Moran statistic, its expected value, variance, a corresponding *z*-score, and a *p*-value. For interpretation and mapping, we classified polygons into the four standard Moran quadrants based on the sign of (*Y_i_* − *Y*^̄^) and of the centered spatial lag Σ*_j_ w_ij_*(*Y_j_* − *Y*^̄^): high–high (hotspots), low–low (coldspots), high–low (high-value outliers among low neighbours), and low–high (low-value outliers among high neighbours) [71]. To account for multiple local tests, we adjusted the resulting local *p*-values using the Benjamini–Hochberg false discovery rate procedure [72, 73], and only polygons with FDR-adjusted *p <* 0.05 were retained as statistically significant LISA clusters in the maps.

#### S1.5 Hospital-level comparison with 2023 CPE surveillance

##### Hospital linkage and surveillance outcome

Hospital-level simulation outputs were linked to 2023 hospital patient-day data and e-SIN CPE surveillance records using FINESS identifiers, standardized as nine-character strings. Hospitals located in departments with codes beginning with 97 or 98 were excluded to remain consistent with the metropolitan-France simulation population. The analysis was restricted to hospitals represented in the simulation and with positive patient-days in 2023. Hospitals that did not appear in the 2023 e-SIN event table were classified as having no recorded CPE case. This classification indicates only that no case was recorded through e-SIN during 2023 and should not be interpreted as evidence that no CPE colonization or transmission occurred.

The primary surveillance outcome was binary:

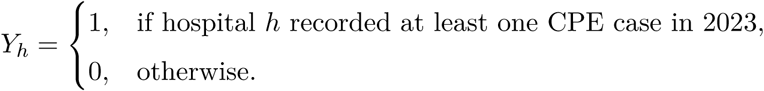

##### Hospital-level simulated relative colonization rate

Patient-level hospital-attributed colonization probabilities were obtained as described in the main-text Methods subsection on aggregation of simulated risk. For each hospital *h*, these probabilities were summed to obtain the expected simulated colonization burden attributed to that hospital, denoted by *A_h_*. Hospital activity was measured using patient-days. Let *PD_h_* denote the total number of days spent by patients in hospital *h* during 2023. The simulated relative colonization rate was defined as

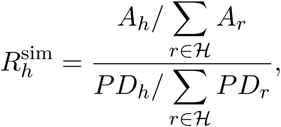

where *H* denotes the set of hospitals included in the hospital-level analysis. A value of 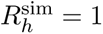 indicates that the hospital’s share of expected simulated colonizations was proportional to its share of patient-days. Values above or below 1 indicate higher or lower simulated colonization rates relative to hospital activity, respectively.

##### Regression analysis

We used multivariable logistic regression to assess whether the simulated relative colonization rate was associated with recording at least one CPE case after accounting for hospital activity:

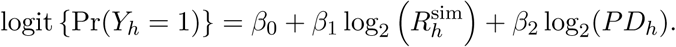

Both predictors were entered as continuous log_2_-transformed variables, so that a one-unit increase on the transformed scale corresponded to a doubling of the original value. Accordingly, exp(*β*_1_) represents the odds ratio associated with a doubling of the simulated relative colonization rate after adjustment for hospital patient-days.

Patient-days were retained as a separate predictor because their role in the regression differs from their role in constructing the relative rate. The relative rate measures simulated colonization in proportion to hospital activity, whereas absolute patient-days reflect the total opportunity for a case to occur, be detected, and be reported. Hospitals with similar relative rates may therefore have different probabilities of recording at least one case if their total activity differs.

A model containing patient-days alone served as the reference model. The additional contribution of the simulated relative colonization rate was evaluated by comparing the full and reference models using a likelihood-ratio test. Two hospitals with a simulated relative colonization rate of zero were excluded because log_2_(0) is undefined, leaving 1,389 hospitals in the fitted model. For visualization in Fig. 3C (in the main paper), adjusted probabilities of recording at least one CPE case were obtained from the fitted model across the observed range of simulated relative colonization rates, with hospital patient-days fixed at their 25th percentile, median, or 75th percentile. Patient-days remained a continuous variable in the fitted regression; these three values were used only to display model-based predictions at representative levels of hospital activity.

#### S1.6 Multivariable beta-regression models

##### Model specification

Tables S5–S6 provide the complete outputs from the multivariable beta-regression models relating simulation-derived colonization risk to patient- and hospital-level characteristics. The conceptual framework and principal findings are described in the main text; here we provide additional details on model estimation, predictor categorization, marginal standardization, and the quantities reported in the supplementary tables. Separate patient- and hospital-level models were fitted using mgcv::bam with a beta-regression likelihood and logit link [62]. The patient-level model was fitted using fast restricted maximum likelihood (method = “fREML”) with discrete = TRUE for computational efficiency on the large patient dataset, whereas the hospital-level model used restricted maximum likelihood (method = “REML”). Because beta regression requires responses strictly within (0, 1), simulated probabilities equal to 0 or 1 were replaced by *ε* or 1 − *ε*, respectively, with *ε* = 10*^−^*^6^. Several continuous predictors were strongly skewed and contained relatively sparse extreme values. These variables were therefore entered as ordered categories rather than imposing a single linear association across their full observed ranges. Category boundaries were defined manually to retain interpretable ranges and resolution across commonly observed values while limiting sparsely populated groups; wider intervals were generally used toward the tails of the distributions. No automated quantile-based binning rule was imposed. Exact category boundaries and the number of observations within each category are reported in Tables S5–S6.

##### Marginal standardization and standardized risk ratios

Adjusted predicted risks were obtained using marginaleffects::avg_predictions() [74]. For each category of a given predictor, that category was assigned to all observations in the corresponding analytic population while all other covariates retained their observed values; predicted colonization probabilities were then averaged over the population. This yielded a population-average adjusted predicted risk, *p̂_c_*, for each category *c*.

The standardized risk ratio was calculated separately within each model as

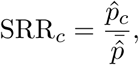

where 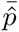 denotes the overall mean model-predicted colonization risk in the same analytic population. Thus, SRR = 1 represents the model-specific average risk, whereas SRR *>* 1 and SRR *<* 1 indicate higher- and lower-than-average adjusted modeled risk, respectively.

Regression coefficients and SRRs represent different comparisons. Coefficients are reported on the logit scale and compare each category with the designated reference category for that predictor, conditional on all other terms in the multivariable model. In contrast, SRRs compare the marginally standardized predicted risk for each category with the overall model-predicted average. Consequently, statistical significance of a model coefficient relative to its predictor-specific reference category does not necessarily imply that the corresponding SRR differs from 1.

##### Table contents

For each predictor category, Tables S5–S6 report the category definition, number and percentage of observations in the analytic sample, regression coefficient on the logit scale relative to the predictor-specific reference category, marginally standardized predicted risk with 95% confidence interval, and corresponding SRR with 95% confidence interval.

##### Patient-level model (Table S5)

The patient-level model included *n* = 5,849,046 patients. Predictors included age, sex, number of admissions, number of wards visited, mean length of stay per ward, number of hospitals visited, mean residence-to-hospital travel distance, predominant ward type, residential population density, and residential socioeconomic deprivation (FDep20). Predominant ward type was defined as the ward type in which each patient accumulated the greatest total length of stay. The multivariable beta-regression model explained 36.4% of the deviance (adjusted *R*^2^ = 0.284).

##### Hospital-level model (Table S6)

The hospital-level model included *n* = 1,389 hospitals with complete information for the outcome and all included covariates. Predictors included hospital mean length of stay, annual admissions, observed hospital capacity, number of medical units, number of ward types, proportion of patients with more than one admission, and inter-hospital connectivity. Observed hospital capacity was defined as the 95th percentile of daily inpatient census in 2023, and inter-hospital connectivity as hospital degree in the patient-sharing network. The model explained 78.5% of the deviance (adjusted *R*^2^ = 0.612).

### S2. Supplementary Figures and Tables

**Figure S1:**
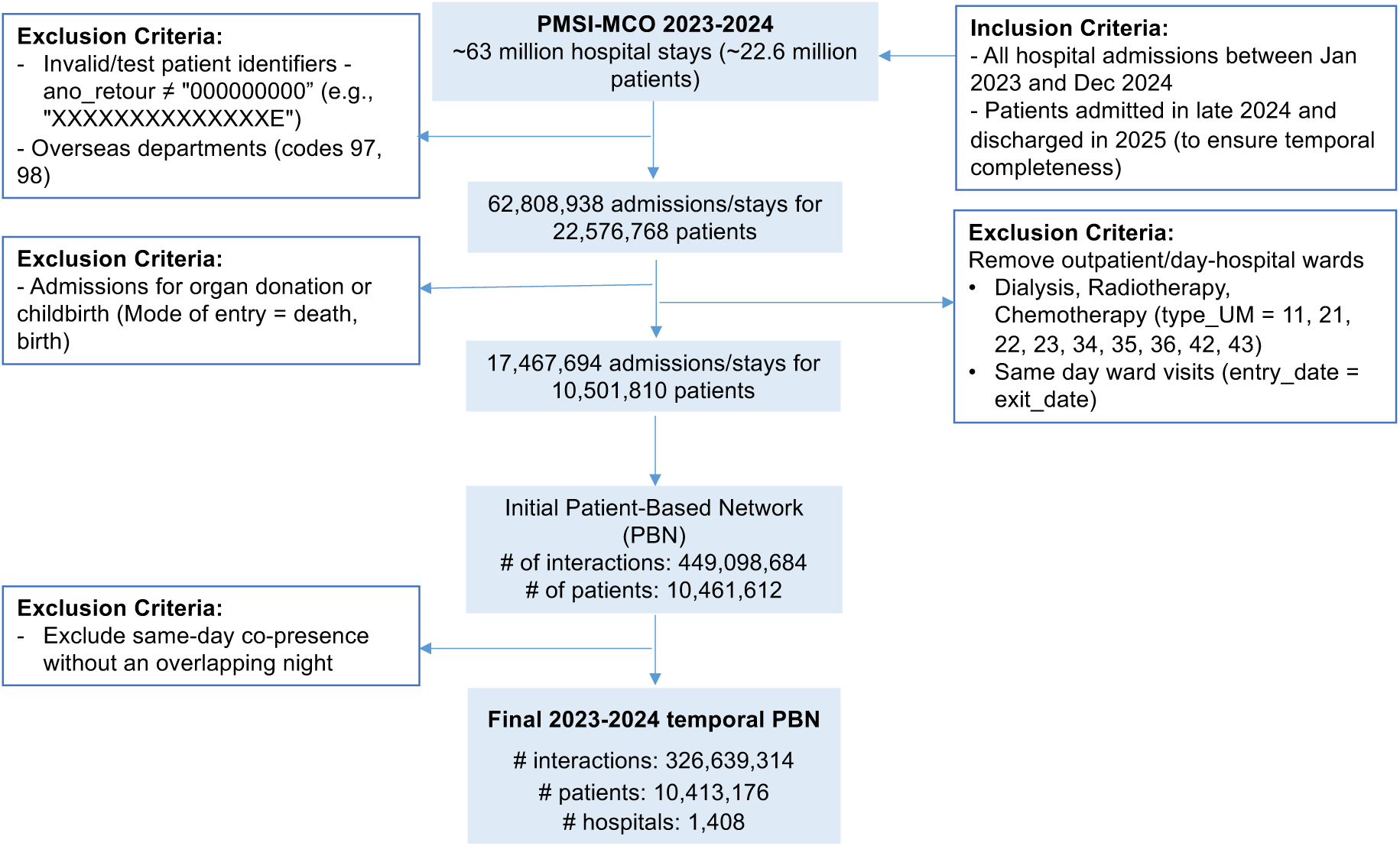
Construction of the 2023–2024 patient-based network (PBN) from the PMSI–MCO database. Flow diagram showing the sequential eligibility criteria applied to PMSI–MCO records from metropolitan France, from all acute-care hospital stays recorded in 2023–2024 to the final temporal PBN used in transmission simulations. Patient trajectories were reconstructed by integrating three PMSI tables: FIXE, containing stay dates, hospital identifiers, patient characteristics, residential codes, and diagnoses; UM, containing movements and lengths of stay within medical units; and IUM, containing medical-unit categories and bed capacity. These tables were linked using stable pseudonymized patient identifiers and unique hospital-stay identifiers. Exclusions comprised invalid or test patient identifiers, overseas hospital sites, inconsistent stay durations, outpatient and day-hospital activity, same-day ward visits, and admissions related to childbirth or organ donation. We also excluded stays related to organ donation or childbirth and records for which the mode of entry was coded as birth or death. In PMSI, each newborn generates a separate medical-unit summary (*résumé d’unité médicale*, RUM), and transfers shortly after birth may produce multiple records with the same admission date but different hospital identifiers or residential codes. RUMs are also generated for stillborn infants after 22 weeks of amenorrhea or with a birth weight ≥ 500 g [75]. These records were excluded because they do not represent conventional inpatient trajectories suitable for reconstructing patient-to-patient transmission pathways. The final patient–patient contacts were defined as co-location in the same medical unit for at least one overlapping night; pairs without an overnight overlap were excluded.

**Figure S2:**
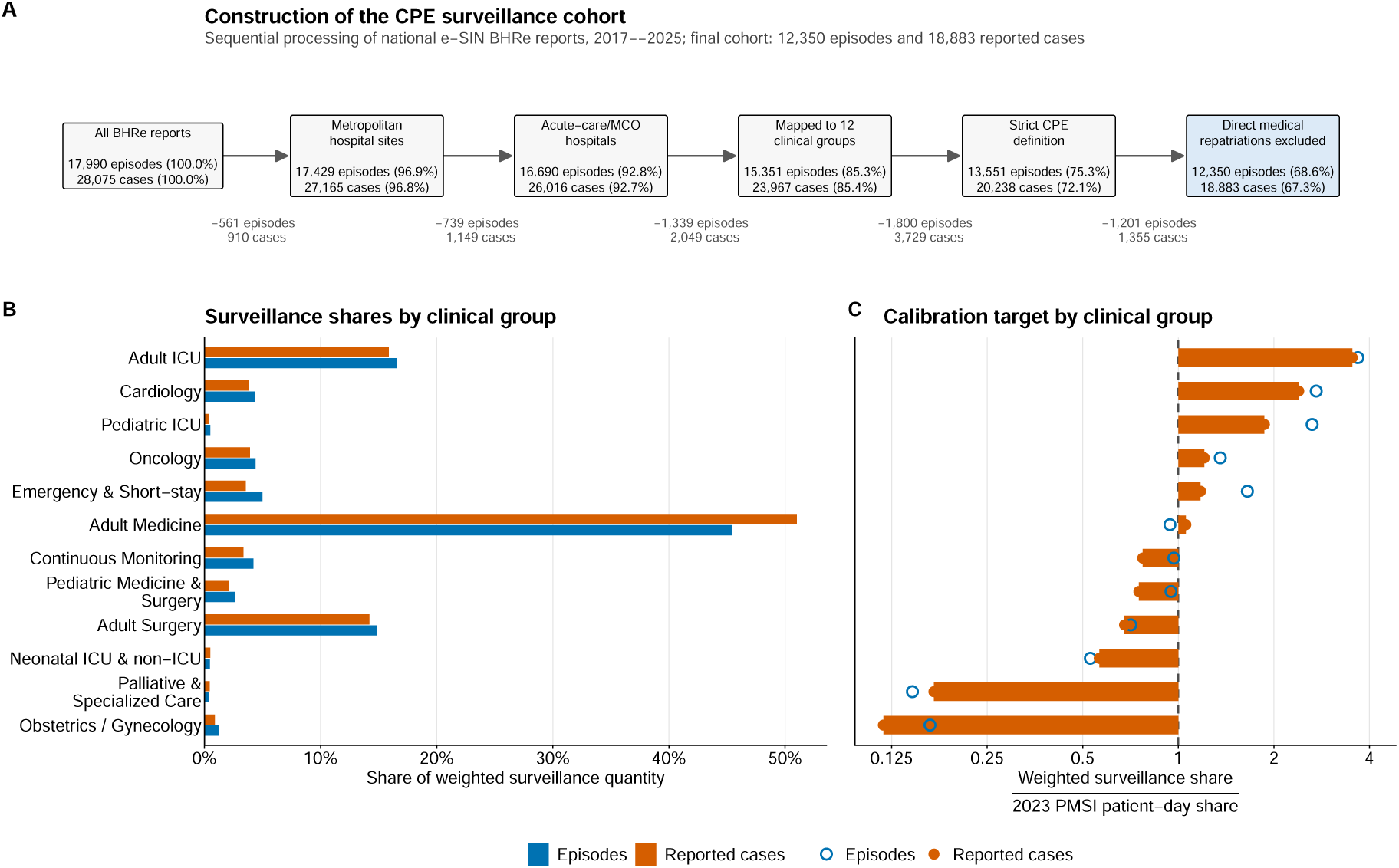
Construction of surveillance-informed clinical-group targets for calibration of CPE transmission. **(A)** Sequential construction of the CPE surveillance cohort from national e-SIN BHRe reports recorded from 2017 through 2025. Values within each box show the number and cumulative percentage of episodes and reported cases retained from the initial dataset; values between boxes show the numbers removed at each filtering step. The final cohort comprised 12,350 CPE episodes and 18,883 reported cases. **(B)** Distribution of weighted surveillance episodes and reported cases across the 12 clinical groups. When an episode included more than one clinical group, its episode weight and reported cases were divided equally among the unique groups represented. Bars show the resulting percentage assigned to each clinical group. **(C)** Clinical-group surveillance shares standardized by the corresponding share of national PMSI patient-days (total number of days spent by patients in each clinical group) in 2023. Orange bars show the reported-case-based ratios used as the primary calibration targets, and open blue circles show the episode-based ratios used as a robustness comparison. The vertical dashed line marks a ratio of 1, corresponding to equal surveillance and patient-day shares. Ratios above 1 indicate greater representation in surveillance than expected from inpatient exposure, whereas ratios below 1 indicate lower representation. The horizontal axis is shown on a log-base-2 scale, with tick labels expressed as the original ratios.

**Figure S3:**
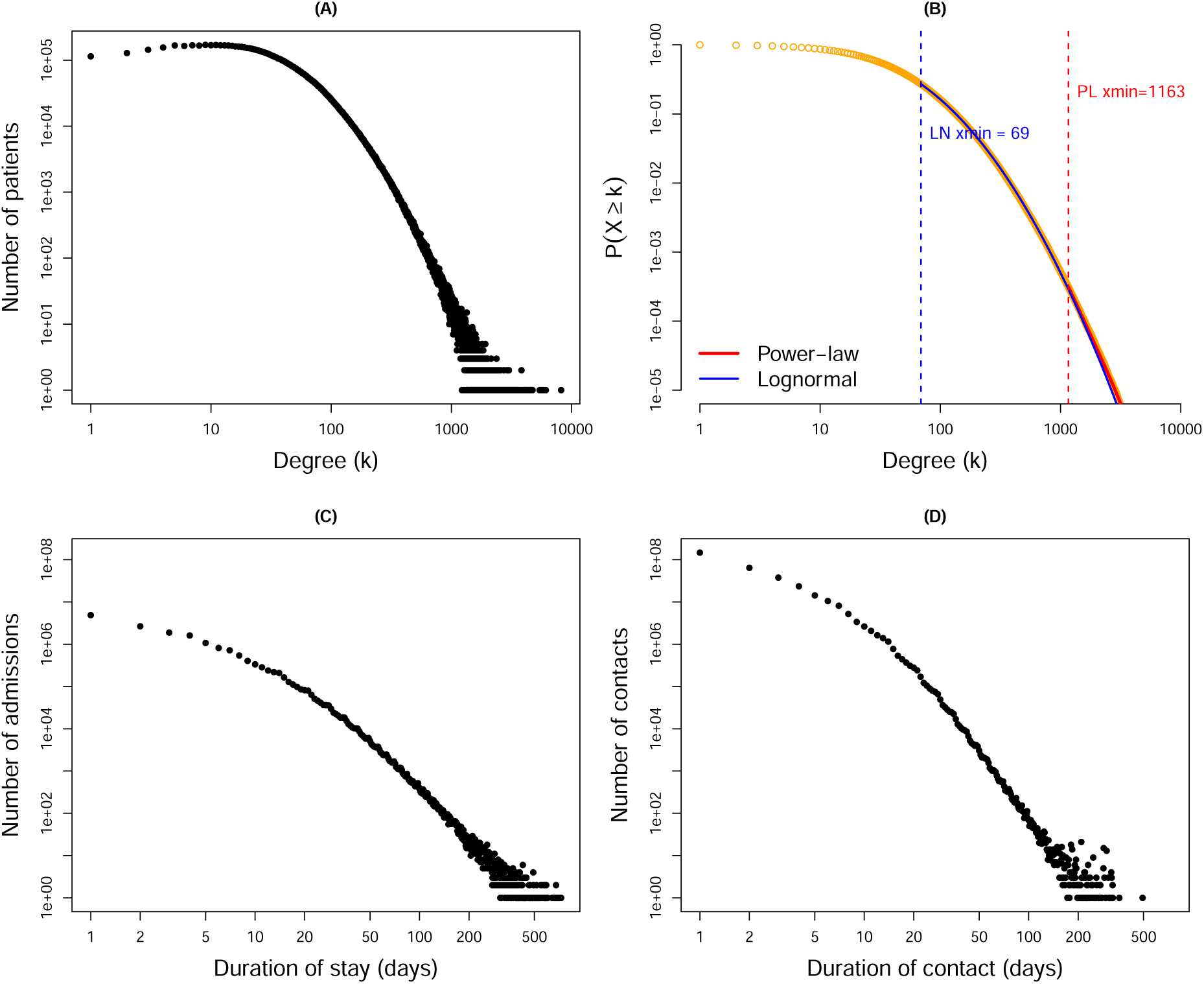
Heterogeneity in the nationwide static PBN-France. All panels use log-log axes and are based on the full 2023–2024 PBN. **(A)** Degree distribution (counts): For each degree value *k*, the y-axis shows the number of patients who shared wards with exactly *k* distinct other patients during the 2023–2024 period. While most patients have few contacts, a small subset are co-located with hundreds to thousands of distinct individuals, producing a strongly heavy-tailed degree distribution. **(B)** Complementary cumulative degree distribution *P* (*X* ≥ *k*), with fitted power-law (red) and lognormal (blue) models. The power-law fit applies only to the extreme upper tail (cutoff *x*_min_ = 1163, covering 3,257 patients, or 0.03% of patients in the PBN), with exponent *γ̂_k_* = 4.73, and thus fails to capture the broader structure of patient connectivity. In contrast, the lognormal fit spans a much wider range of degrees (starting at *x*_min_ = 69, including 2,816,646 patients, or 27.05% of patients in the PBN), encompassing both moderately and highly connected patients. Together, panels (A)–(B) show that the PBN is heavy-tailed but not scale-free [76]—suggesting that transmission may propagate through a broad spectrum of patients rather than relying solely on extreme “super-spreaders.” **(C)** Length of stay distribution: Distribution of hospitalization duration (days) across 17,467,694 admissions represented in the 2023–2024 PBN, after restricting durations to values compatible with the two-year observation window. While most stays are short, a substantial fraction extend over several weeks or months. **(D)** Contact duration distribution: Distribution of contact duration between patient pairs, defined as the number of overlapping ward-days, based on 326,639,314 pairwise temporal co-location events. Most contacts are brief, but some persist for long periods. Together, these distributions show substantial structural heterogeneity in patient connectivity, length of stay, and contact duration, all of which are explicitly represented in the transmission simulations.

**Figure S4:**
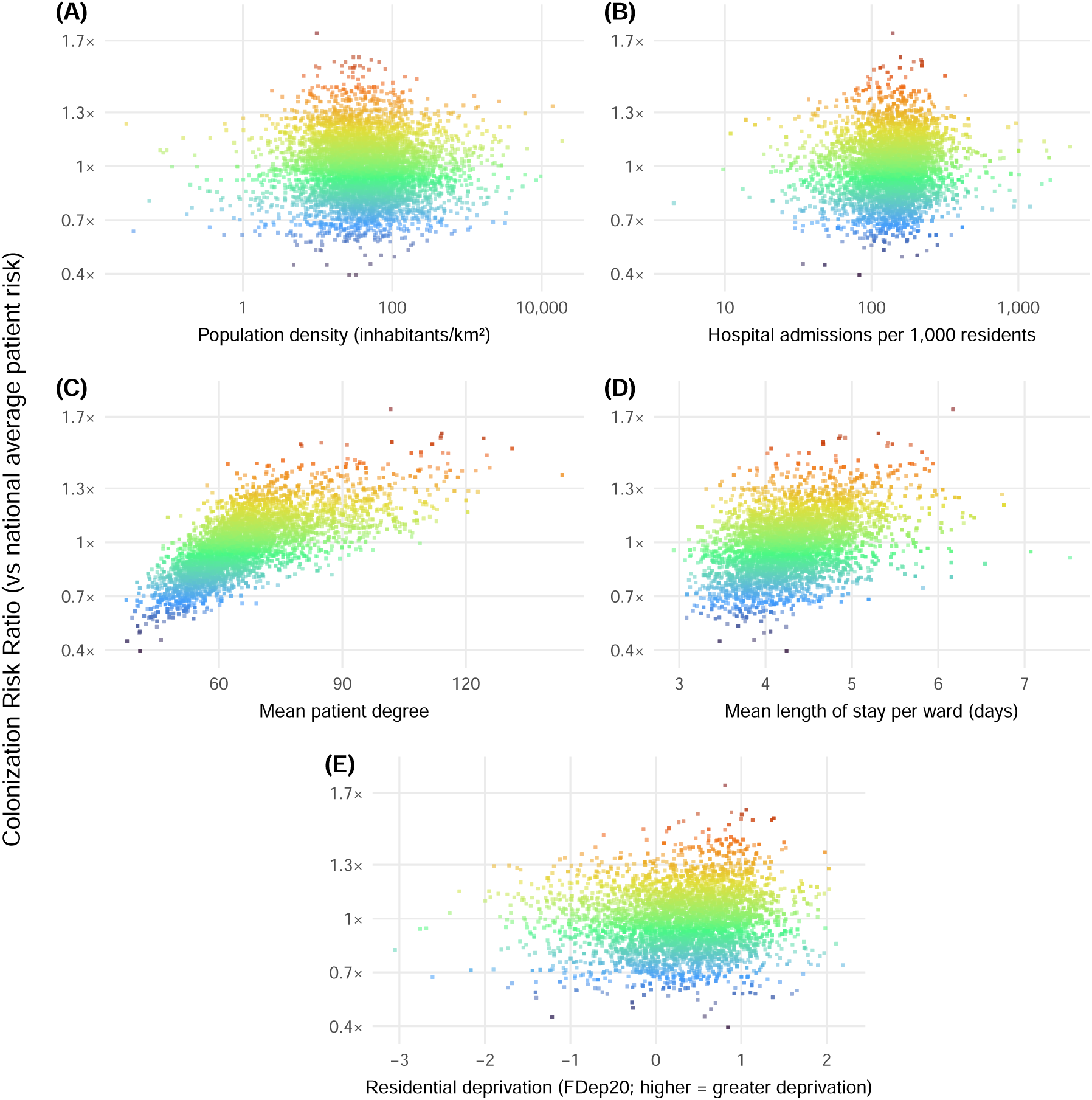
Additional descriptive correlates of simulated residential colonization risk supporting Fig. 1C–D of the main text. Each point corresponds to one population-density grid cell, as in Fig. 1C of the main text, and is colored according to the residential colonization risk ratio relative to the national mean simulated patient risk. **(A)** Residential colonization risk ratio across the continuous range of population density, complementing the population-density categories shown in Fig. 1D. **(B)** Residential colonization risk ratio according to hospital admissions per 1,000 residents, representing an area-level measure of hospital use. **(C)** Residential colonization risk ratio according to mean patient degree in the patient-based network. **(D)** Residential colonization risk ratio according to mean length of stay per ward. **(E)** Residential colonization risk ratio according to residential socioeconomic deprivation measured using the French Deprivation Index (FDep20), with higher values indicating greater deprivation. Patient degree and mean length of stay per ward were calculated at the individual-patient level and then averaged within residential areas. Residential-area summaries were assigned to the population-density grid cells and, where multiple residential areas contributed to a grid cell, combined using the number of PBN patients as weights. Together, the panels provide complementary descriptive views of how simulated residential colonization risk varies with population context, hospital use, patient-network connectivity, duration of hospital exposure, and residential socioeconomic deprivation. Population density in panel **A** and hospital admissions per 1,000 residents in panel **B** are shown on logarithmic x-axes.

**Figure S5:**
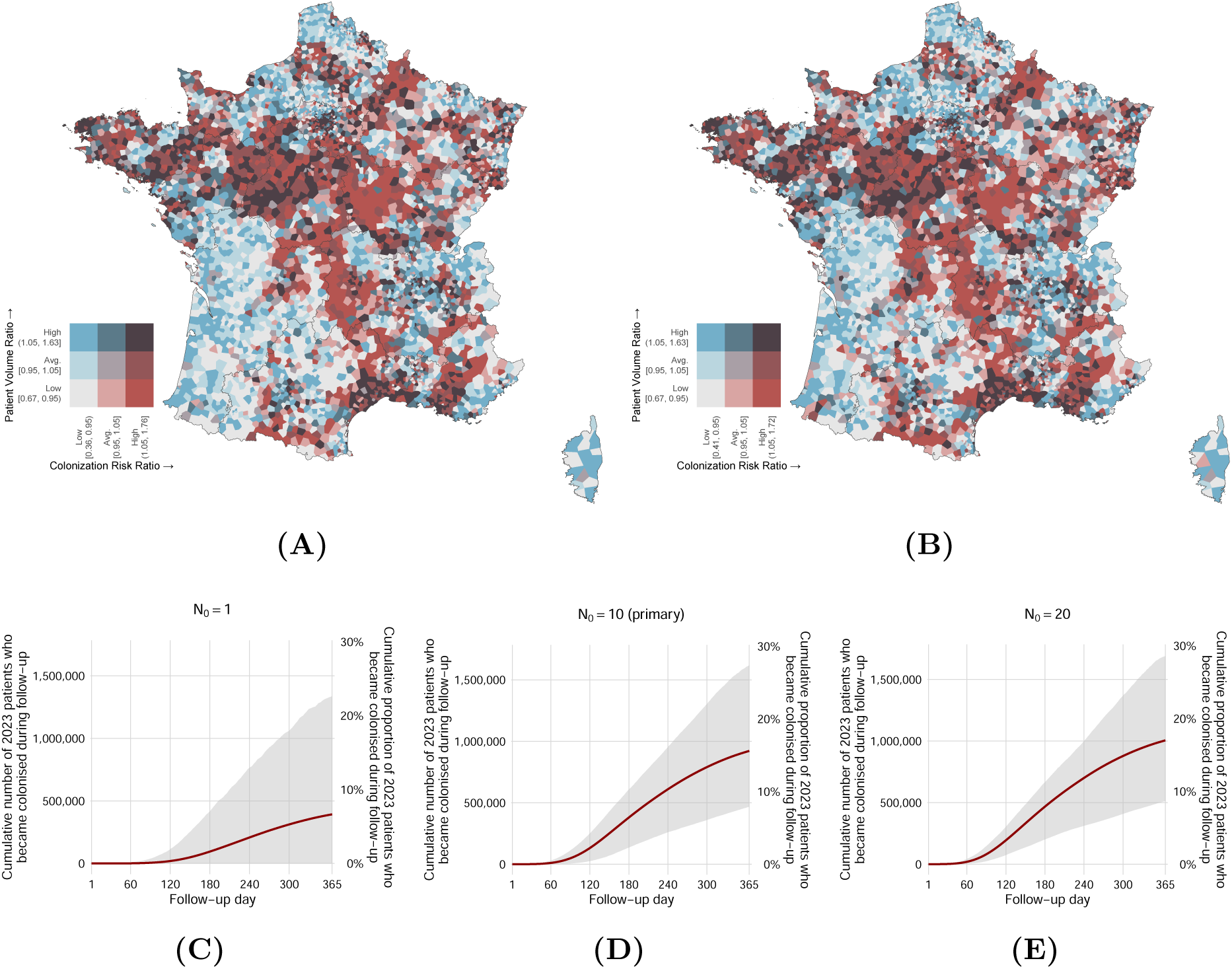
Sensitivity of residential colonization-risk patterns and cumulative colonization burden to the number of initially colonized patients. **(A,B)** Residential colonization-risk maps obtained with *N*_0_ = 1 (A) and *N*_0_ = 20 (B) initially colonized patients; the corresponding *N*_0_ = 10 map is presented in the primary analysis. Each Voronoi polygon represents a residential unit, and its color jointly indicates the simulated colonization-risk ratio and patient-volume ratio relative to their respective national means. **(C–E)** Cumulative number and proportion of patients present in 2023 who became colonized at least once during the 365-day follow-up under *N*_0_ = 1 (C), *N*_0_ = 10 (D; primary scenario), and *N*_0_ = 20 (E). Solid lines show the mean across simulations, and shaded ribbons show the 2.5th–97.5th percentile range across simulations. The left *y*-axis reports the cumulative number of patients, whereas the right *y*-axis expresses the same quantity as a proportion of the 5,910,857-patient 2023 target population. Initial seed colonizations were excluded from this outcome. All scenarios used the primary *D* = 180-day colonization duration and the same calibrated transmission parameters, with only the number of initially colonized patients varied. The primary *N*_0_ = 10 scenario included 2,000 simulations, whereas the *N*_0_ = 1 and *N*_0_ = 20 sensitivity scenarios included 1,000 simulations each. By day 365, the mean cumulative proportion of patients who had become colonized was 6.65% for *N*_0_ = 1, 15.60% for *N*_0_ = 10, and 17.01% for *N*_0_ = 20; the corresponding 2.5th–97.5^th^ percentile ranges were 0–22.61%, 7.93–27.36%, and 8.67–28.58%, respectively. Patient-level colonization probabilities remained strongly concordant with the primary *N*_0_ = 10 scenario, with Spearman rank correlations of *ρ* = 0.973 for *N*_0_ = 1 and *ρ* = 0.990 for *N*_0_ = 20 (Pearson *r* = 0.993 and 0.998, respectively). Thus, varying the initial condition affected the magnitude of simulated colonization burden more strongly than the relative patient-level and residential distribution of risk. Because all transmission parameters were held fixed, these comparisons specifically assess sensitivity to the number of initial colonization introductions.

**Figure S6:**
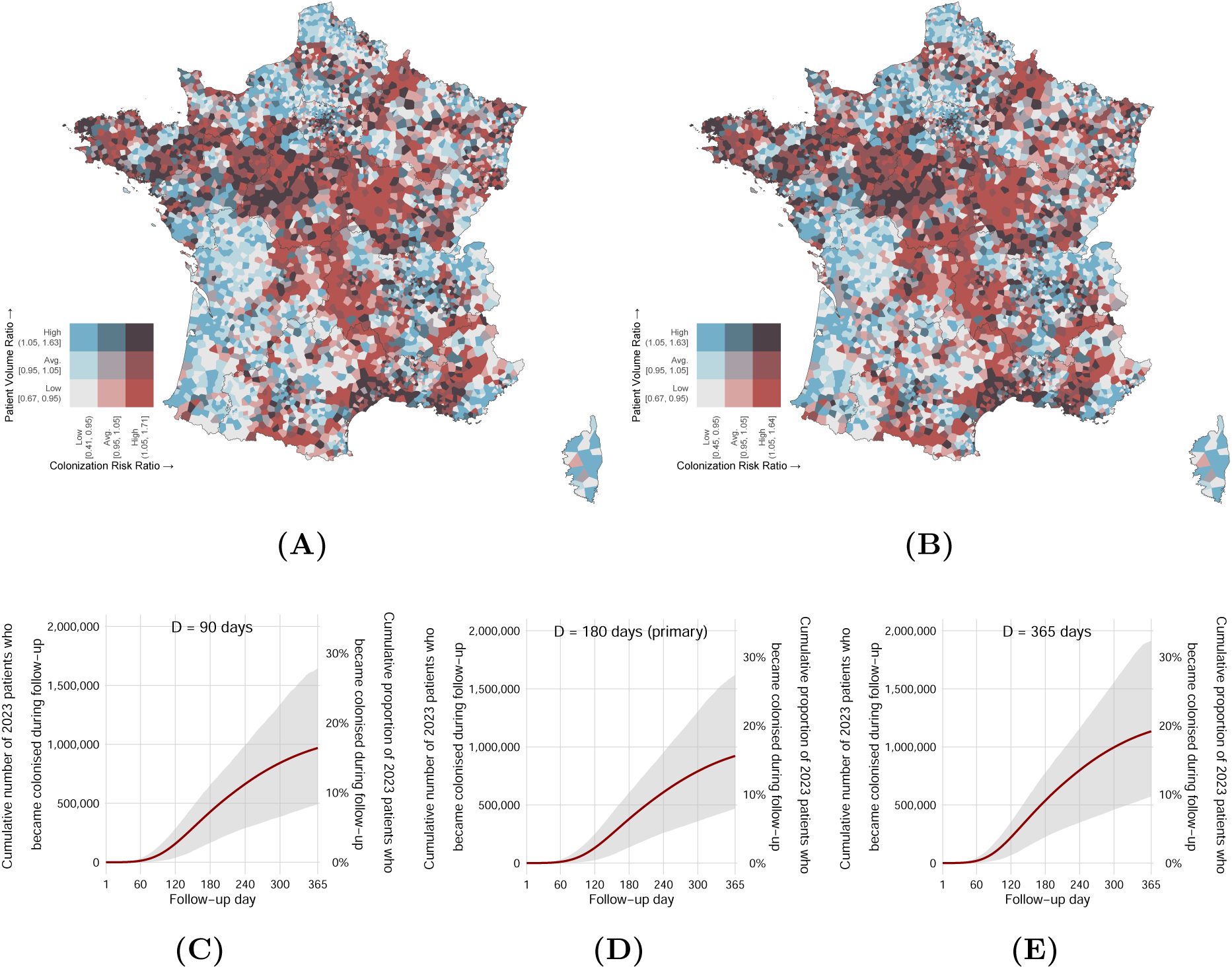
Sensitivity of residential colonization-risk patterns and cumulative colonization burden to the assumed duration of colonization. **(A,B)** Residential colonization-risk maps obtained under assumed colonization durations of *D* = 90 days (A) and *D* = 365 days (B); the corresponding *D* = 180-day map is presented in the primary analysis. Each Voronoi polygon represents a residential unit, and its color jointly indicates the simulated colonization-risk ratio and patient-volume ratio relative to their respective national means. **(C–E)** Cumulative number and proportion of patients present in 2023 who became colonized at least once during the 365-day follow-up under *D* = 90 days (C), *D* = 180 days (D; primary scenario), and *D* = 365 days (E). Solid lines show the mean across simulations, and shaded ribbons show the 2.5th–97.5th percentile range across simulations. The left *y*-axis reports the cumulative number of patients, whereas the right *y*-axis expresses the same quantity as a proportion of the 5,910,857-patient 2023 target population. For each assumed duration, the clinical-group-specific transmission multipliers were recalibrated independently using the same surveillance-informed calibration procedure. The primary *D* = 180-day scenario included 2,000 simulations, whereas the *D* = 90- and *D* = 365-day sensitivity scenarios included 1,000 simulations each. Patient-level colonization probabilities remained highly concordant with those from the primary *D* = 180-day scenario, with Spearman rank correlations of *ρ* = 0.987 for *D* = 90 days and *ρ* = 0.988 for *D* = 365 days across all 5,910,857 patients. Thus, although the absolute probability scale changed somewhat, particularly under *D* = 365, the relative ordering of patients and the broad residential risk pattern were largely preserved. Because the transmission multipliers were recalibrated separately for each duration, these comparisons assess the robustness of the complete surveillance-informed modeling framework rather than the isolated effect of changing colonization duration while holding transmission parameters fixed.

**Figure S7:**
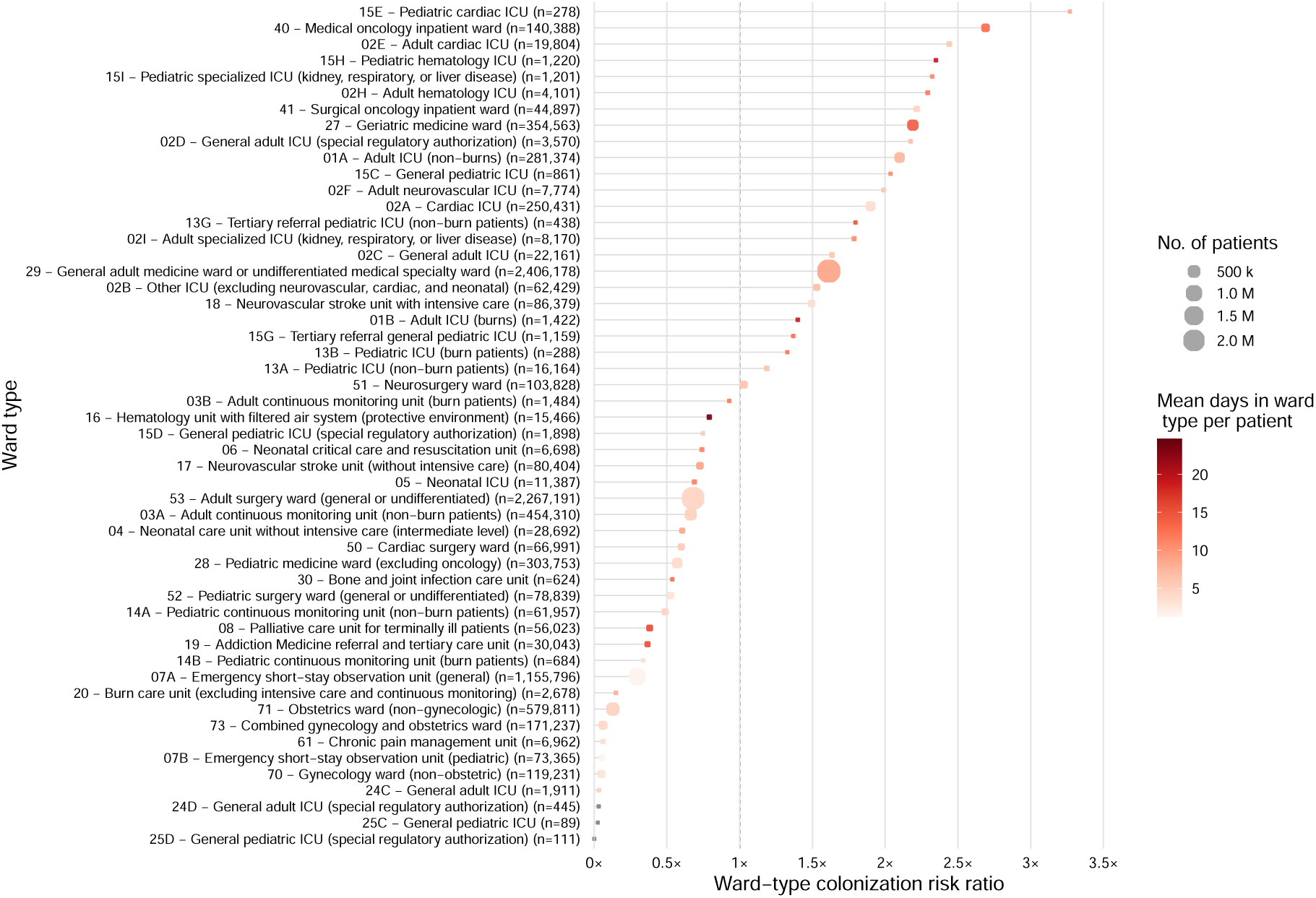
Variation in simulated colonization risk and duration of patient exposure across hospital ward types. Each bubble represents one PMSI ward type (type_UM) included in the detailed ward-type analysis. The horizontal position shows the ward-type colonization risk ratio, calculated from the mean patient-level colonization probability attributed to that ward type relative to the national reference mean across patient–ward-type observations. The vertical dashed line at 1 marks this national reference; values above or below 1 therefore indicate higher or lower simulated ward-associated colonization risk, respectively. Bubble size represents the number of unique patients contributing to each ward type. Bubble color represents the mean number of days spent in that ward type per patient in 2023. Lighter colors therefore indicate shorter mean duration of exposure and darker colors indicate longer mean duration. PMSI ward-type codes, descriptive labels, and the corresponding numbers of patients are shown on the *y*-axis. The figure illustrates substantial heterogeneity in simulated colonization risk across ward types with widely differing patient volumes and durations of inpatient exposure.

**Table S4:** Mapping of PMSI ward types to clinical groups and calibrated transmission parameters used in the primary analysis. Ward types are defined according to the PMSI ward-type classification (type_UM). For each clinical group, the table reports the included ward-type codes, the number of ward types represented, the number of unique patients who received care in at least one ward type belonging to that group, and the calibrated transmission multiplier *w_g_*. Values in brackets give the corresponding daily per-colonized-neighbor transmission probability, *β_g_*= *β*_0_*w_g_*, with *β*_0_ = 0.05. For the primary analysis with a mean colonization duration of 180 days, calibrated multipliers ranged from *w_g_* = 0.100 to 4.467, corresponding to *β_g_* = 0.0050 to 0.2233. These multipliers are surveillance-informed clinical-setting modifiers and should not be interpreted as direct biological estimates of CPE transmissibility. Detailed ward-type labels and corresponding simulated colonization risks are shown in Fig. S7.

| Clinical group | PMSI ward-type (type_UM) codes | No. of ward types | Patients, $n$ | $w_g$ [$\beta_g$] |
| --- | --- | --- | --- | --- |
| Adult Medicine | 17, 27, 29, 61 | 4 | 2,613,524 | 0.316 [0.0158] |
| Adult Surgery | 50, 51, 53 | 3 | 2,395,287 | 0.237 [0.0118] |
| Obstetrics / Gynecology | 70, 71, 73 | 3 | 825,024 | 0.158 [0.0079] |
| Oncology | 40, 41 | 2 | 173,920 | 1.997 [0.0999] |
| Palliative & Specialized Care | 08, 16, 19, 20, 30 | 5 | 104,119 | 0.100 [0.0050] |
| Cardiology | 02A, 02E, 15E | 3 | 266,286 | 3.361 [0.1681] |
| Adult ICU | 01A, 01B, 02B, 02C, 02D, 02F, 02H, 02I, 18, 24C, 24D | 11 | 441,681 | 4.000 [0.2000] |
| Pediatric Medicine & Surgery | 28, 52 | 2 | 367,230 | 0.793 [0.0396] |
| Pediatric ICU | 13A, 13B, 13G, 15C, 15D, 15G, 15H, 15I, 25C, 25D | 10 | 21,704 | 1.641 [0.0821] |
| Continuous Monitoring | 03A, 03B, 14A, 14B | 4 | 517,385 | 0.708 [0.0354] |
| Neonatal ICU & non-ICU | 04, 05, 06 | 3 | 36,876 | 4.467 [0.2233] |
| Emergency & Short-stay | 07A, 07B | 2 | 1,228,240 | 0.904 [0.0452] |

**Figure S8:**
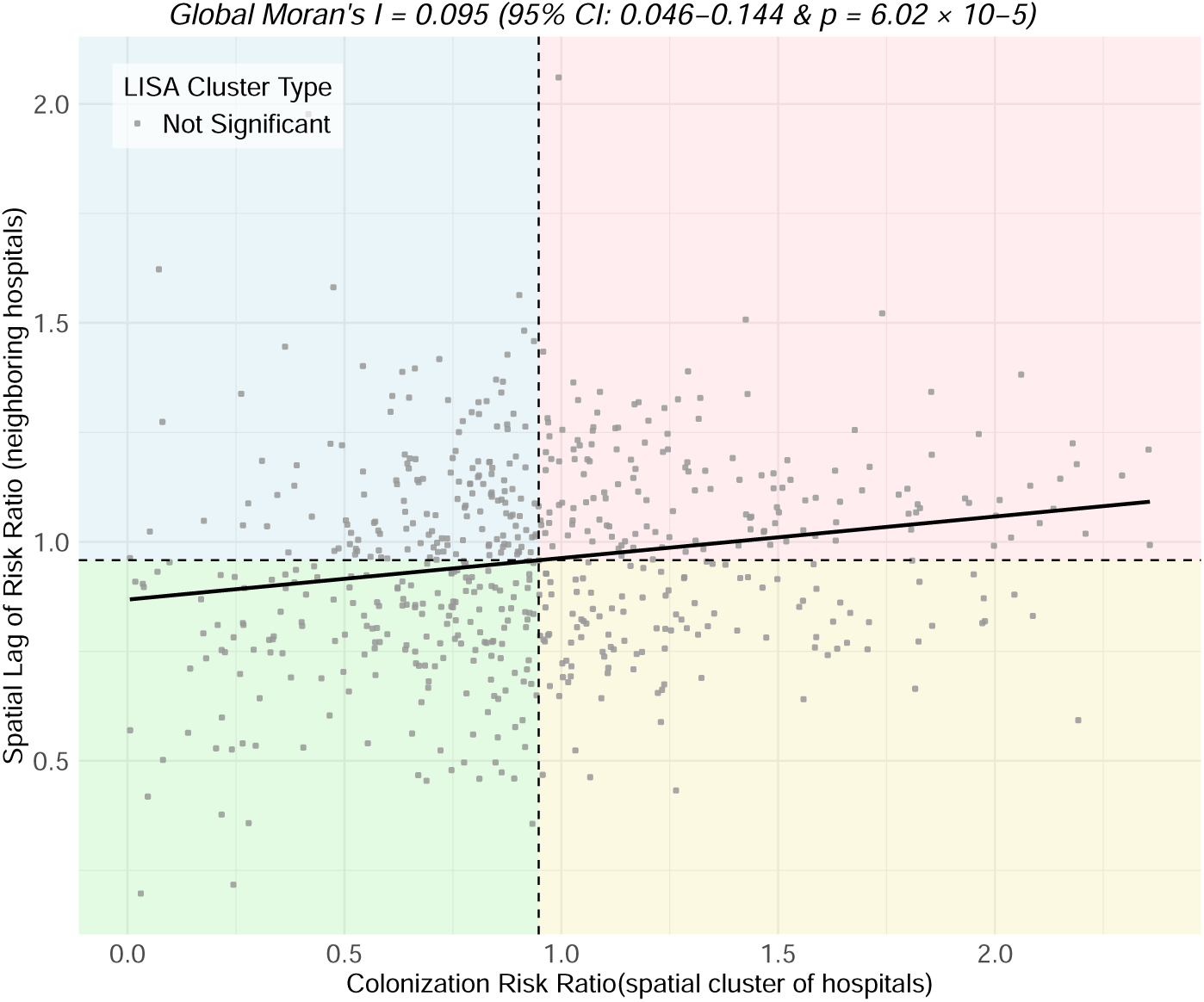
Spatial autocorrelation of hospital-level simulated colonization risk. The Moran scatterplot shows the colonization risk ratio for each hospital-level Voronoi polygon on the horizontal axis and its spatial lag on the vertical axis, calculated as the row-standardized mean risk ratio of neighboring polygons defined by queen contiguity. Dashed vertical and horizontal lines indicate the corresponding global means and divide the scatterplot into the four conventional Moran quadrants: high–high, low–low, high–low, and low–high. The solid line is the least-squares regression line, whose slope corresponds to Global Moran’s *I*. Hospital-level simulated colonization risk showed weak statistically significant positive spatial autocorrelation (Global Moran’s *I* = 0.095, approximate 95% CI: 0.046–0.144; *P <* 0.001). Local Indicators of Spatial Association (LISA) identified no individual polygons that remained statistically significant after correction for multiple testing; points are therefore shown uniformly in gray.

**Table S5:**
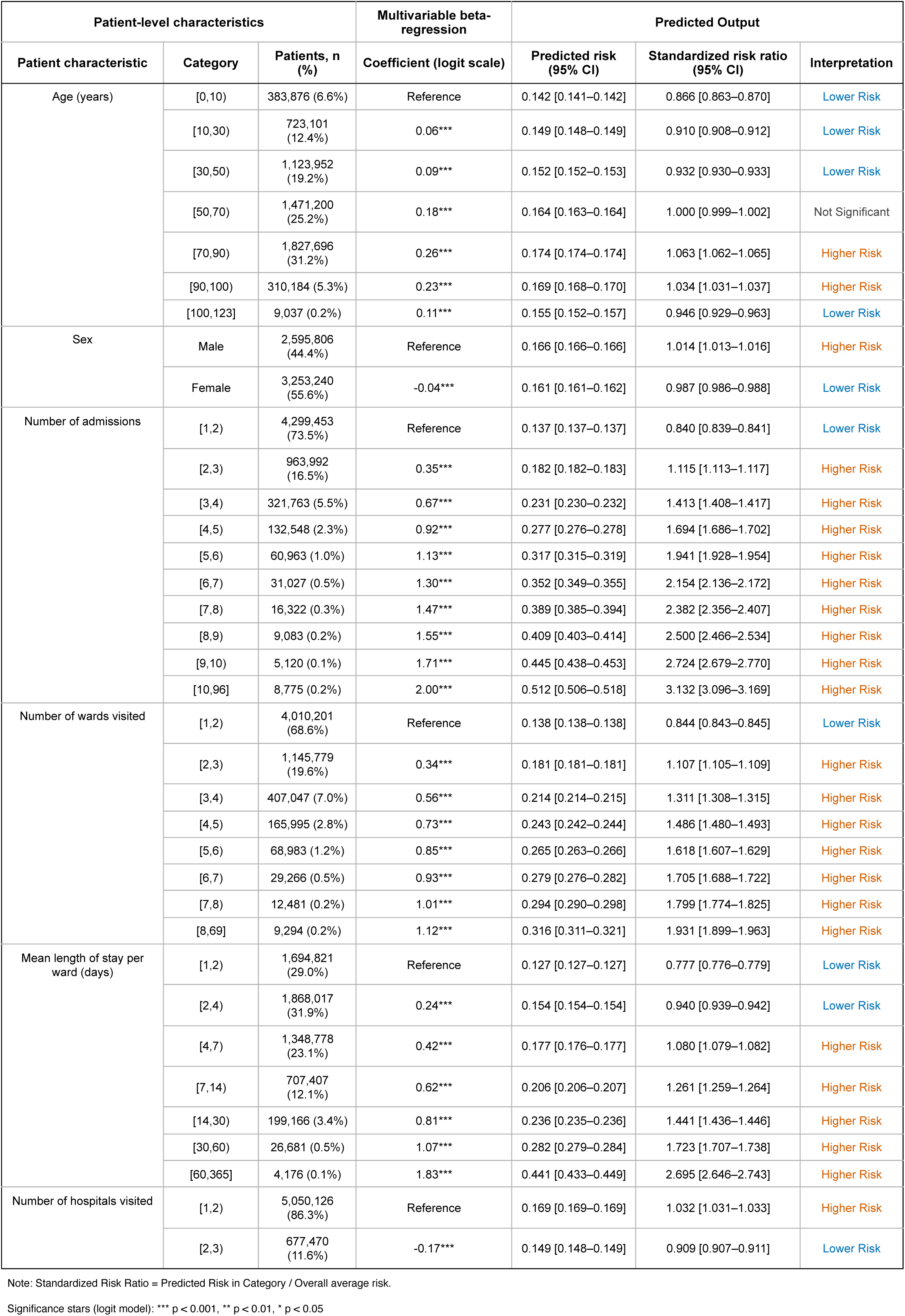
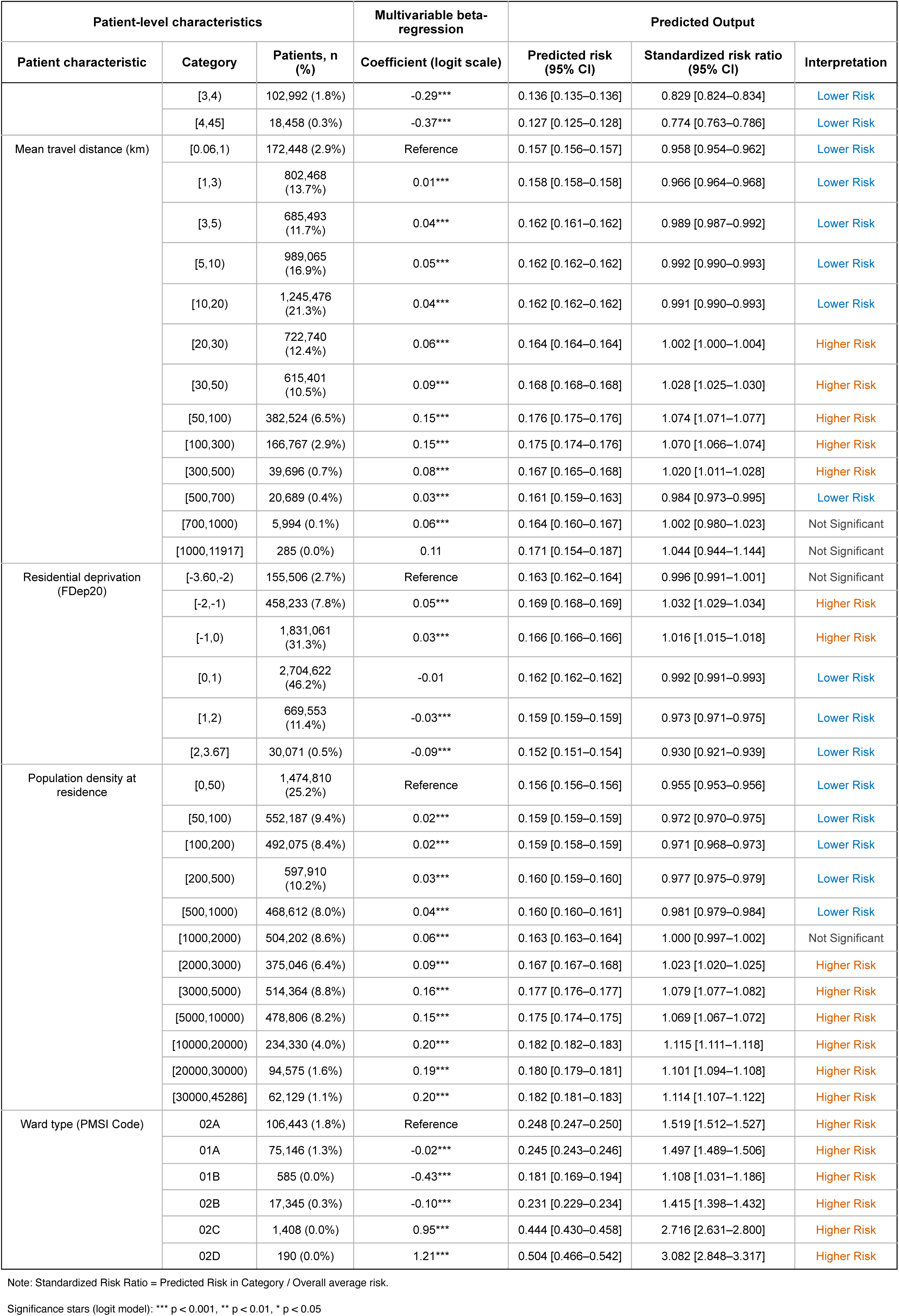
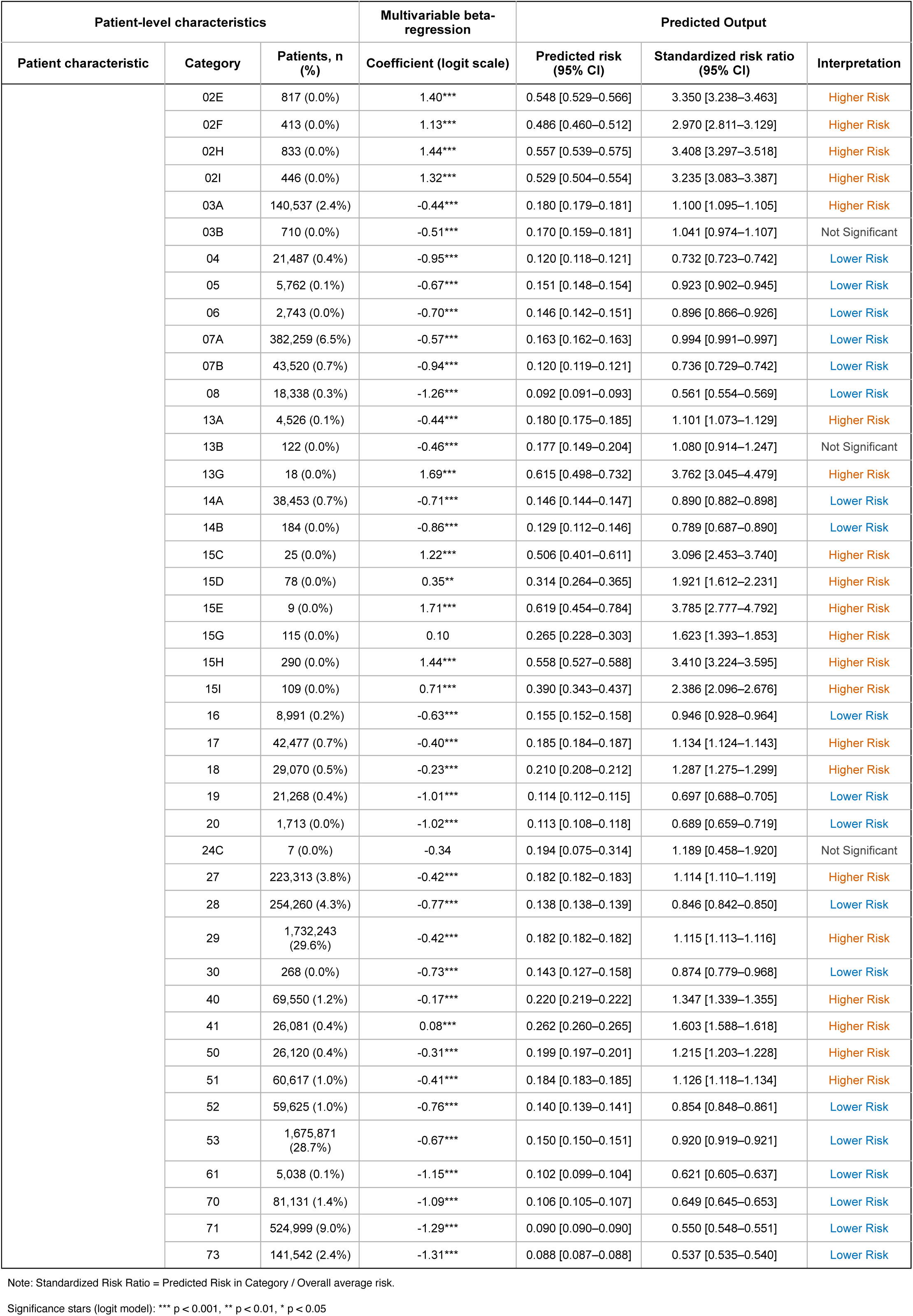
Patient-level multivariable beta-regression results. Complete category-specific estimates from the model of simulated patient-level colonization probability. Adjusted predicted risks were obtained by marginal standardization, and standardized risk ratios (SRRs) compare each category-specific adjusted predicted risk with the overall model-predicted average. SRR = 1 denotes the overall modeled average; SRR *>* 1 and SRR *<* 1 indicate higher- and lower-than-average modeled risk, respectively. Regression-coefficient significance refers to comparisons with the predictor-specific reference category: *** *P <* 0.001, ** *P <* 0.01, * *P <* 0.05.

**Table S6:** Hospital-level multivariable beta-regression results. Complete category-specific estimates from the model of simulated hospital-level colonization probability across 1,389 hospitals. Adjusted predicted risks were obtained by marginal standardization, and standardized risk ratios (SRRs) compare each category-specific adjusted predicted risk with the overall model-predicted average. SRR = 1 denotes the overall modeled average; SRR *>* 1 and SRR *<* 1 indicate higher- and lower-than-average modeled risk, respectively. Regression-coefficient significance refers to comparisons with the predictor-specific reference category: *** *P <* 0.001, ** *P <* 0.01, * *P <* 0.05.

| Hospital-Level Characteristics |  |  | Multivariable beta-regression model | Predicted Output |  |  |
| --- | --- | --- | --- | --- | --- | --- |
| Hospital categorical variables | Category | No. Hospitals (%) | Coefficient (logit scale) | Predicted risk [95% CI] | Standardized risk ratio [95% CI] | Interpretation |
| Mean length of stay (days) | [0,5) | 335 (24.1%) | Reference | 0.033 [0.030 – 0.036] | 0.323 [0.293 – 0.354] | Lower Risk |
|  | [5,10) | 674 (48.5%) | 1.22*** | 0.102 [0.098 – 0.106] | 1.001 [0.965 – 1.038] | Not Significant |
|  | [10,15) | 209 (15.0%) | 1.69*** | 0.152 [0.142 – 0.162] | 1.490 [1.389 – 1.590] | Higher Risk |
|  | [15,20) | 111 (8.0%) | 2.05*** | 0.202 [0.183 – 0.220] | 1.978 [1.796 – 2.160] | Higher Risk |
|  | [20,25) | 46 (3.3%) | 2.13*** | 0.215 [0.186 – 0.243] | 2.105 [1.826 – 2.384] | Higher Risk |
|  | [25,48.7] | 16 (1.2%) | 2.47*** | 0.274 [0.223 – 0.324] | 2.682 [2.183 – 3.181] | Higher Risk |
| Inter-hospital connectivity | [0,20) | 147 (10.4%) | Reference | 0.091 [0.078 – 0.104] | 0.891 [0.766 – 1.016] | Not Significant |
|  | [20,40) | 274 (19.5%) | 0.01 | 0.091 [0.083 – 0.100] | 0.896 [0.815 – 0.977] | Lower Risk |
|  | [40,60) | 159 (11.3%) | 0.07 | 0.097 [0.089 – 0.105] | 0.951 [0.869 – 1.034] | Not Significant |
|  | [60,80) | 143 (10.2%) | 0.03 | 0.093 [0.085 – 0.102] | 0.916 [0.836 – 0.997] | Lower Risk |
|  | [80,100) | 128 (9.1%) | 0.08 | 0.098 [0.089 – 0.107] | 0.958 [0.871 – 1.046] | Not Significant |
|  | [100,150) | 196 (13.9%) | 0.15 | 0.103 [0.096 – 0.111] | 1.012 [0.938 – 1.085] | Not Significant |
|  | [150,200) | 117 (8.3%) | 0.20 | 0.108 [0.098 – 0.117] | 1.056 [0.961 – 1.151] | Not Significant |
|  | [200,300) | 146 (10.4%) | 0.29* | 0.117 [0.106 – 0.128] | 1.144 [1.036 – 1.253] | Higher Risk |
|  | [300,400) | 63 (4.5%) | 0.34* | 0.122 [0.105 – 0.139] | 1.196 [1.030 – 1.361] | Higher Risk |
|  | [400,690] | 35 (2.5%) | 0.42** | 0.130 [0.107 – 0.153] | 1.275 [1.052 – 1.498] | Higher Risk |
| Observed hospital capacity | [0,25) | 434 (31.2%) | Reference | 0.061 [0.054 – 0.068] | 0.598 [0.528 – 0.669] | Lower Risk |
|  | [25,50) | 240 (17.3%) | 0.45*** | 0.091 [0.083 – 0.099] | 0.892 [0.813 – 0.971] | Lower Risk |
|  | [50,100) | 270 (19.4%) | 0.74*** | 0.117 [0.107 – 0.127] | 1.150 [1.051 – 1.249] | Higher Risk |
|  | [100,200) | 211 (15.2%) | 0.92*** | 0.135 [0.120 – 0.151] | 1.326 [1.175 – 1.477] | Higher Risk |
|  | [200,300) | 100 (7.2%) | 0.96*** | 0.140 [0.116 – 0.163] | 1.370 [1.142 – 1.597] | Higher Risk |
|  | [300,500) | 82 (5.9%) | 1.06*** | 0.152 [0.121 – 0.182] | 1.490 [1.191 – 1.788] | Higher Risk |
|  | [500,700) | 36 (2.6%) | 1.29*** | 0.181 [0.135 – 0.227] | 1.776 [1.325 – 2.227] | Higher Risk |
|  | [700,1068] | 18 (1.3%) | 1.37*** | 0.194 [0.108 – 0.279] | 1.900 [1.062 – 2.738] | Higher Risk |
| Annual admissions | [0,200) | 110 (7.9%) | Reference | 0.040 [0.032 – 0.049] | 0.396 [0.311 – 0.481] | Lower Risk |
|  | [200,500) | 174 (12.5%) | 0.61*** | 0.071 [0.059 – 0.082] | 0.692 [0.580 – 0.805] | Lower Risk |
|  | [500,1000) | 143 (10.3%) | 0.91*** | 0.092 [0.081 – 0.104] | 0.904 [0.792 – 1.015] | Not Significant |
|  | [1000,2000) | 184 (13.2%) | 1.05*** | 0.104 [0.094 – 0.114] | 1.024 [0.925 – 1.122] | Not Significant |
|  | [2000,3000) | 107 (7.7%) | 1.01*** | 0.101 [0.089 – 0.113] | 0.987 [0.870 – 1.104] | Not Significant |
|  | [3000,5000) | 172 (12.4%) | 1.13*** | 0.112 [0.099 – 0.125] | 1.098 [0.970 – 1.225] | Not Significant |
|  | [5000,10000) | 212 (15.2%) | 1.24*** | 0.122 [0.107 – 0.138] | 1.201 [1.045 – 1.358] | Higher Risk |
|  | [10000,20000) | 192 (13.8%) | 1.49*** | 0.150 [0.126 – 0.173] | 1.466 [1.239 – 1.693] | Higher Risk |
|  | [20000,40000) | 80 (5.8%) | 1.46*** | 0.146 [0.114 – 0.179] | 1.433 [1.115 – 1.752] | Higher Risk |
|  | [40000,58594] | 17 (1.2%) | 1.51*** | 0.152 [0.085 – 0.220] | 1.495 [0.835 – 2.154] | Not Significant |
| Number of ward types | [0,2) | 405 (29.1%) | Reference | 0.134 [0.119 – 0.148] | 1.311 [1.171 – 1.450] | Higher Risk |
|  | [2,4) | 332 (23.9%) | -0.25*** | 0.108 [0.099 – 0.118] | 1.064 [0.970 – 1.158] | Not Significant |
|  | [4,6) | 191 (13.7%) | -0.30*** | 0.104 [0.095 – 0.112] | 1.017 [0.931 – 1.102] | Not Significant |
|  | [6,10) | 202 (14.5%) | -0.46*** | 0.091 [0.083 – 0.099] | 0.892 [0.817 – 0.967] | Lower Risk |

Note: Standardized Risk Ratio = Predicted Risk in Category / Overall average risk.

Significance stars (logit model): \*\*\* p < 0.001, \*\* p < 0.01, \* p < 0.05
| Hospital-Level Characteristics |  |  | Multivariable beta-regression model | Predicted Output |  |  |
| --- | --- | --- | --- | --- | --- | --- |
| Hospital categorical variables | Category | No. Hospitals (%) | Coefficient (logit scale) | Predicted risk [95% CI] | Standardized risk ratio [95% CI] | Interpretation |
|  | [10,15) | 128 (9.2%) | -0.57*** | 0.082 [0.072 – 0.092] | 0.808 [0.710 – 0.906] | Lower Risk |
|  | [15,20) | 83 (6.0%) | -0.60*** | 0.080 [0.068 – 0.092] | 0.784 [0.663 – 0.906] | Lower Risk |
|  | [20,25) | 30 (2.2%) | -0.57*** | 0.082 [0.065 – 0.099] | 0.804 [0.636 – 0.972] | Lower Risk |
|  | [25,32] | 20 (1.4%) | -0.62*** | 0.078 [0.058 – 0.099] | 0.770 [0.570 – 0.969] | Lower Risk |
| Number of medical units | [0,5) | 553 (39.8%) | Reference | 0.148 [0.134 – 0.162] | 1.454 [1.316 – 1.591] | Higher Risk |
|  | [5,10) | 275 (19.8%) | -0.32*** | 0.114 [0.105 – 0.123] | 1.119 [1.028 – 1.210] | Higher Risk |
|  | [10,20) | 239 (17.2%) | -0.44*** | 0.104 [0.096 – 0.112] | 1.020 [0.943 – 1.097] | Not Significant |
|  | [20,30) | 97 (7.0%) | -0.76*** | 0.079 [0.071 – 0.087] | 0.775 [0.694 – 0.855] | Lower Risk |
|  | [30,50) | 111 (8.0%) | -0.92*** | 0.068 [0.060 – 0.076] | 0.667 [0.588 – 0.747] | Lower Risk |
|  | [50,100) | 96 (6.9%) | -1.01*** | 0.063 [0.053 – 0.073] | 0.616 [0.518 – 0.714] | Lower Risk |
|  | [100,168] | 20 (1.4%) | -1.19*** | 0.054 [0.037 – 0.070] | 0.526 [0.362 – 0.691] | Lower Risk |
| % of patients with > 1 admission | [0,5) | 96 (6.8%) | Reference | 0.033 [0.027 – 0.039] | 0.328 [0.269 – 0.387] | Lower Risk |
|  | [5,10) | 235 (16.7%) | 0.75*** | 0.068 [0.062 – 0.073] | 0.665 [0.609 – 0.720] | Lower Risk |
|  | [10,15) | 387 (27.5%) | 1.15*** | 0.097 [0.092 – 0.101] | 0.949 [0.903 – 0.994] | Lower Risk |
|  | [15,20) | 475 (33.7%) | 1.30*** | 0.110 [0.105 – 0.114] | 1.076 [1.034 – 1.118] | Higher Risk |
|  | [20,25) | 155 (11.0%) | 1.39*** | 0.119 [0.111 – 0.127] | 1.168 [1.092 – 1.245] | Higher Risk |
|  | [25,35) | 38 (2.7%) | 1.77*** | 0.163 [0.146 – 0.181] | 1.600 [1.427 – 1.773] | Higher Risk |
|  | [35,73.93] | 22 (1.6%) | 1.92*** | 0.183 [0.158 – 0.209] | 1.799 [1.546 – 2.052] | Higher Risk |

Note: Standardized Risk Ratio = Predicted Risk in Category / Overall average risk.

## References

[1] Mohsen Naghavi, Stein Emil Vollset, Kevin S Ikuta, Lucien R Swetschinski, Authia P Gray, Eve E Wool, Gisela Robles Aguilar, Tomislav Mestrovic, Georgia Smith, Chieh Han, et al. Global burden of bacterial antimicrobial resistance 1990–2021: a systematic analysis with forecasts to 2050. The Lancet, 404(10459):1199–1226, 2024.

[2] Christopher JL Murray, Kevin Shunji Ikuta, Fablina Sharara, Lucien Swetschinski, Gisela Robles Aguilar, Authia Gray, Chieh Han, Catherine Bisignano, Puja Rao, Eve Wool, et al. Global burden of bacterial antimicrobial resistance in 2019: a systematic analysis. The lancet, 399(10325):629–655, 2022.

[3] M. J. Bonten, M. K. Hayden, C. Nathan, J. van Voorhis, M. Matushek, S. Slaughter, T. Rice, and R. A. Weinstein. Epidemiology of colonisation of patients and environment with vancomycin-resistant enterococci. Lancet (London, England), 348(9042):1615–1619, December 1996. ISSN 0140-6736. doi: 10.1016/S0140-6736(96)02331-8.

[4] Sandra S Richter and Dror Marchaim. Screening for carbapenem-resistant enterobacteriaceae: who, when, and how? Virulence, 8(4):417–426, 2017.

[5] MM MacKinnon and KD Allen. Long-term mrsa carriage in hospital patients. Journal of Hospital Infection, 46(3):216–221, 2000.

[6] Frederic S Zimmerman, Marc V Assous, Tali Bdolah-Abram, Tamar Lachish, Amos M Yinnon, and Yonit Wiener-Well. Duration of carriage of carbapenem-resistant enterobacteriaceae following hospital discharge. American journal of infection control, 41(3):190–194, 2013.

[7] Agnes Scanvic, Ljiljiana Denic, Stéphanie Gaillon, Pascal Giry, Antoine Andremont, and Jean-Christophe Lucet. Duration of colonization by methicillin-resistant staphylococcus aureus after hospital discharge and risk factors for prolonged carriage. Clinical Infectious Diseases, 32(10):1393–1398, 2001.

[8] David L. Smith, Jonathan Dushoff, Eli N. Perencevich, Anthony D. Harris, and Simon A. Levin. Persistent colonization and the spread of antibiotic resistance in nosocomial pathogens: Resistance is a regional problem. Proceedings of the National Academy of Sciences, 101 (10):3709–3714, March 2004. doi: 10.1073/pnas.0400456101. Publisher: Proceedings of the National Academy of Sciences.

[9] M. J. Bonten, S. Slaughter, A. W. Ambergen, M. K. Hayden, J. van Voorhis, C. Nathan, and R. A. Weinstein. The role of “colonization pressure” in the spread of vancomycin-resistant enterococci: an important infection control variable. Archives of Internal Medicine, 158 (10):1127–1132, May 1998. ISSN 0003-9926. doi: 10.1001/archinte.158.10.1127.

[10] World Health Organization. Global report on infection prevention and control. Global report, World Health Organization, Geneva, 2024. URL https://www.who.int/publications/i/item/9789240090991.

[11] Kidu Gidey, Meles Tekie Gidey, Berhane Yohannes Hailu, Zigbey Brhane Gebreamlak, and Yirga Legesse Niriayo. Clinical and economic burden of healthcare-associated infections: A prospective cohort study. Plos one, 18(2):e0282141, 2023.

[12] Lindsey M. Weiner, Amy K. Webb, Brandi Limbago, Margaret A. Dudeck, Jean Patel, Alexander J. Kallen, Jonathan R. Edwards, and Dawn M. Sievert. Antimicrobial-Resistant Pathogens Associated With Healthcare-Associated Infections: Summary of Data Reported to the National Healthcare Safety Network at the Centers for Disease Control and Prevention, 2011–2014. Infection control and hospital epidemiology, 37(11):1288–1301, November 2016. ISSN 0899-823X. doi: 10.1017/ice.2016.174.

[13] Alex W. Friedrich. Control of hospital acquired infections and antimicrobial resistance in Europe: the way to go. Wiener Medizinische Wochenschrift, 169(1):25–30, February 2019. ISSN 1563-258X. doi: 10.1007/s10354-018-0676-5.

[14] Shelley S Magill, Erin O’Leary, Sarah J Janelle, Deborah L Thompson, Ghinwa Dumyati, Joelle Nadle, Lucy E Wilson, Marion A Kainer, Ruth Lynfield, Samantha Greissman, et al. Changes in prevalence of health care–associated infections in us hospitals. New England Journal of Medicine, 379(18):1732–1744, 2018.

[15] Derek Cocker, Gabriel Birgand, Nina Zhu, Jesus Rodriguez-Manzano, Raheelah Ahmad, Kondwani Jambo, Anna S. Levin, and Alison Holmes. Healthcare as a driver, reservoir and amplifier of antimicrobial resistance: opportunities for interventions. Nature Reviews Micro-biology, 22(10):636–649, October 2024. ISSN 1740-1534. doi: 10.1038/s41579-024-01076-4. Publisher: Nature Publishing Group.

[16] Ondrej Zahornacký, Štefan Porubčin, Alena Rovňáková, and Pavol Jarčuška. Gram-negative rods on inanimate surfaces of selected hospital facilities and their nosocomial significance. International Journal of Environmental Research and Public Health, 19(10):6039, 2022. doi: 10.3390/ijerph19106039.

[17] Po Ying Chia, Sharmila Sengupta, Anjanna Kukreja, Sasheela S.L. Ponnampalavanar, Oon Tek Ng, and Kalisvar Marimuthu. The role of hospital environment in transmissions of multidrug-resistant gram-negative organisms. Antimicrobial Resistance & Infection Control, 9(1):29, February 2020. ISSN 2047-2994. doi: 10.1186/s13756-020-0685-1.

[18] Anita Bhalla, Nicole J. Pultz, Delores M. Gries, Amy J. Ray, Elizabeth C. Eckstein, David C. Aron, and Curtis J. Donskey. Acquisition of Nosocomial Pathogens on Hands After Contact With Environmental Surfaces Near Hospitalized Patients. Infection Control & Hospital Epidemiology, 25(2):164–167, February 2004. ISSN 0899-823X, 1559-6834. doi: 10.1086/502369.

[19] Anna L. Costa, Gaetano Pierpaolo Privitera, Giorgio Tulli, and Giulio Toccafondi. Infection Prevention and Control. In Liam Donaldson, Walter Ricciardi, Susan Sheridan, and Riccardo Tartaglia, editors, Textbook of Patient Safety and Clinical Risk Management. Springer, Cham (CH), 2021. ISBN 978-3-030-59402-2 978-3-030-59403-9. URL http://www.ncbi.nlm.nih.gov/books/NBK585600/.

[20] Hanjue Xia, Johannes Horn, Monika J Piotrowska, Konrad Sakowski, André Karch, Hannan Tahir, Mirjam Kretzschmar, and Rafael Mikolajczyk. Effects of incomplete inter-hospital network data on the assessment of transmission dynamics of hospital-acquired infections. PLoS computational biology, 17(5):e1008941, 2021.

[21] Tjibbe Donker, Jacco Wallinga, and Hajo Grundmann. Patient referral patterns and the spread of hospital-acquired infections through national health care networks. PLoS computational biology, 6(3):e1000715, 2010.

[22] Monika J Piotrowska, Konrad Sakowski, André Karch, Hannan Tahir, Johannes Horn, Mirjam E Kretzschmar, and Rafael T Mikolajczyk. Modelling pathogen spread in a healthcare network: Indirect patient movements. PLOS Computational Biology, 16(11): e1008442, 2020.

[23] Teresa Ita, Ulzii-Orshikh Luvsansharav, Rachel M. Smith, Robert Mugoh, Charchil Ayodo, Beatrice Oduor, Moureen Jepleting, Walter Oguta, Caroline Ouma, Jane Juma, Godfrey Bigogo, Samuel Kariuki, Brooke M. Ramay, Mark Caudell, Clayton Onyango, Linus Ndegwa, Jennifer R. Verani, Susan Bollinger, Aditya Sharma, Guy H. Palmer, Douglas R. Call, and Sylvia Omulo. Prevalence of colonization with multidrug-resistant bacteria in communities and hospitals in Kenya. Scientific Reports, 12(1):22290, December 2022. ISSN 2045-2322. doi: 10.1038/s41598-022-26842-3. Publisher: Nature Publishing Group.

[24] BS Cooper, GF Medley, SP Stone, CC Kibbler, BD Cookson, JA Roberts, G Duckworth, R Lai, and S Ebrahim. Methicillin-resistant staphylococcus aureus in hospitals and the community: stealth dynamics and control catastrophes. Proceedings of the National Academy of Sciences, 101(27):10223–10228, 2004.

[25] Teresa M. Coque, Rafael Cantón, Ana Elena Pérez-Cobas, Miguel D. Fernández-de Bobadilla, and Fernando Baquero. Antimicrobial Resistance in the Global Health Network: Known Unknowns and Challenges for Efficient Responses in the 21st Century. Microorganisms, 11 (4):1050, April 2023. ISSN 2076-2607. doi: 10.3390/microorganisms11041050.

[26] H. Grundmann and B. Hellriegel. Mathematical modelling: a tool for hospital infection control. The Lancet. Infectious Diseases, 6(1):39–45, January 2006. ISSN 1473-3099. doi: 10.1016/S1473-3099(05)70325-X.

[27] Marc J. M Bonten, Daren J Austin, and Marc Lipsitch. Understanding the Spread of Antibiotic Resistant Pathogens in Hospitals: Mathematical Models as Tools for Control. Clinical infectious diseases: an official publication of the Infectious Diseases Society of America, 33:1739–46, December 2001. doi: 10.1086/323761.

[28] Emily Kajita, Justin T. Okano, Erin N. Bodine, Scott P. Layne, and Sally Blower. Modelling an outbreak of an emerging pathogen. Nature Reviews. Microbiology, 5(9):700–709, September 2007. ISSN 1740-1534. doi: 10.1038/nrmicro1660.

[29] Bruce Y. Lee, Sarah M. McGlone, Yeohan Song, Taliser R. Avery, Stephen Eubank, Chung-Chou Chang, Rachel R. Bailey, Diane K. Wagener, Donald S. Burke, Richard Platt, and Susan S. Huang. Social Network Analysis of Patient Sharing Among Hospitals in Orange County, California. American Journal of Public Health, 101(4):707–713, April 2011. ISSN 0090-0036. doi: 10.2105/AJPH.2010.202754. Publisher: American Public Health Association.

[30] Rania Assab, Narimane Nekkab, Pascal Crepey, Pascal Astagneau, Didier Guillemot, Lulla Opatowski, and Laura Temime. Mathematical models of infection transmission in healthcare settings: recent advances from the use of network structured data. Current Opinion in Infectious Diseases, 30(4):410–418, 2017.

[31] Lulla Opatowski, Didier Guillemot, Pierre-Yves Boëlle, and Laura Temime. Contribution of mathematical modeling to the fight against bacterial antibiotic resistance. Current opinion in infectious diseases, 24(3):279–287, 2011.

[32] Gwenan M Knight, Nicholas G Davies, Caroline Colijn, Francesc Coll, Tjibbe Donker, Danna R Gifford, Rebecca E Glover, Mark Jit, Elizabeth Klemm, Sonja Lehtinen, et al. Mathematical modelling for antibiotic resistance control policy: do we know enough? BMC infectious diseases, 19(1):1011, 2019.

[33] Sen Pei, Flaviano Morone, Fredrik Liljeros, Hernan Makse, and Jeffrey L Shaman. Inference and control of the nosocomial transmission of methicillin-resistant staphylococcus aureus. Elife, 7:e40977, 2018.

[34] Bruce Y. Lee, Sarah M. McGlone, Kim F. Wong, S. Levent Yilmaz, Taliser R. Avery, Yeohan Song, Richard Christie, Stephen Eubank, Shawn T. Brown, Joshua M. Epstein, Jon I. Parker, Donald S. Burke, Richard Platt, and Susan S. Huang. Modeling the Spread of Methicillin-Resistant Staphylococcus aureus (MRSA) Outbreaks throughout the Hospitals in Orange County, California. Infection Control and Hospital Epidemiology, 32(6):562–572, June 2011. ISSN 0899-823X. doi: 10.1086/660014.

[35] Jacob E. Simmering, Linnea A. Polgreen, David R. Campbell, Joseph E. Cavanaugh, and Philip M. Polgreen. Hospital Transfer Network Structure as a Risk Factor for *Clostridium difficile* Infection. Infection Control & Hospital Epidemiology, 36(9):1031–1037, September 2015. ISSN 0899-823X, 1559-6834. doi: 10.1017/ice.2015.130.

[36] Juan Fernández-Gracia, Jukka-Pekka Onnela, Michael L. Barnett, Víctor M. Eguíluz, and Nicholas A. Christakis. Influence of a patient transfer network of US inpatient facilities on the incidence of nosocomial infections. Scientific Reports, 7(1):2930, June 2017. ISSN 2045-2322. doi: 10.1038/s41598-017-02245-7.

[37] Michael J Ray, Michael Y Lin, Angela S Tang, M Allison Arwady, Mary Alice Lavin, Erica Runningdeer, Dejan Jovanov, and William E Trick. Regional Spread of an Outbreak of Carbapenem-Resistant Enterobacteriaceae Through an Ego Network of Healthcare Facilities. Clinical Infectious Diseases, 67(3):407–410, July 2018. ISSN 1058-4838, 1537-6591. doi: 10.1093/cid/ciy084.

[38] Tjibbe Donker, Hajo Grundmann, Laura Temime, Pascal Crépey, Patrizio Pezzotti, Julie V Robotham, Gerolf de Boer, Petra Fadgyas-Freyler, Francesco Di Ruscio, Alex W Friedrich, et al. Towards a europe-wide reconstruction and analysis of hospital networks. Clinical Microbiology and Infection, 2026.

[39] Tjibbe Donker, Jacco Wallinga, Richard Slack, and Hajo Grundmann. Hospital networks and the dispersal of hospital-acquired pathogens by patient transfer. PloS one, 7(4):e35002, 2012.

[40] Narimane Nekkab, Pascal Astagneau, Laura Temime, and Pascal Crepey. Spread of hospital-acquired infections: A comparison of healthcare networks. PLoS computational biology, 13 (8):e1005666, 2017.

[41] Mariano Ciccolini, Tjibbe Donker, Hajo Grundmann, Marc JM Bonten, and Mark EJ Woolhouse. Efficient surveillance for healthcare-associated infections spreading between hospitals. Proceedings of the National Academy of Sciences, 111(6):2271–2276, 2014.

[42] Hannan Tahir, Luis Eduardo López-Cortés, Axel Kola, Dafna Yahav, André Karch, Hanjue Xia, Johannes Horn, Konrad Sakowski, Monika J Piotrowska, Leonard Leibovici, et al. Relevance of intra-hospital patient movements for the spread of healthcare-associated infections within hospitals-a mathematical modeling study. PLoS computational biology, 17 (2):e1008600, 2021.

[43] Birgitte Freiesleben de Blasio and Gianpaolo Scalia Tomba. How do patients move within the norwegian hospital system? a comprehensive ward-and hospital-level network analysis. medRxiv, pages 2026–07, 2026.

[44] Sen Pei, Dwayne Seeram, Seth Blumberg, Bo Shopsin, Anne-Catrin Uhlemann, and Jeffrey Shaman. Inferring asymptomatic carriers of antimicrobial-resistant organisms in hospitals using genomic, microbiological and patient mobility data. Nature Communications, 16(1): 10140, 2025.

[45] Sen Pei, Fredrik Liljeros, and Jeffrey Shaman. Identifying asymptomatic spreaders of antimicrobial-resistant pathogens in hospital settings. Proceedings of the National Academy of Sciences, 118(37), September 2021. ISSN 0027-8424, 1091-6490. doi: 10.1073/pnas.2111190118. Publisher: Proceedings of the National Academy of Sciences.

[46] Ashleigh Myall, James R Price, Robert L Peach, Mohamed Abbas, Sid Mookerjee, Nina Zhu, Isa Ahmad, Damien Ming, Farzan Ramzan, Daniel Teixeira, Christophe Graf, Andrea Y Weiße, Stephan Harbarth, Alison Holmes, and Mauricio Barahona. Prediction of hospital-onset COVID-19 infections using dynamic networks of patient contact: an international retrospective cohort study. The Lancet. Digital Health, 4(8):e573–e583, August 2022. ISSN 2589-7500. doi: 10.1016/S2589-7500(22)00093-0.

[47] Racha Gouareb, Alban Bornet, Dimitrios Proios, Sónia Gonçalves Pereira, and Douglas Teodoro. Detection of patients at risk of multidrug-resistant enterobacteriaceae infection using graph neural networks: a retrospective study. Health Data Science, 3:0099, 2023.

[48] Kaniz Fatema Madhobi, Ananth Kalyanaraman, Deverick J. Anderson, Elizabeth Dodds Ashley, Rebekah W. Moehring, and Eric T. Lofgren. Use of Contact Networks to Estimate Potential Pathogen Risk Exposure in Hospitals. JAMA Network Open, 5(8):e2225508, August 2022. ISSN 2574-3805. doi: 10.1001/jamanetworkopen.2022.25508.

[49] Audrey Duval, Thomas Obadia, Pierre-Yves Boëlle, Eric Fleury, Jean-Louis Herrmann, Didier Guillemot, Laura Temime, Lulla Opatowski, and i Bird Study group. Close proximity interactions support transmission of esbl-k. pneumoniae but not esbl-e. coli in healthcare settings. PLoS computational biology, 15(5):e1006496, 2019.

[50] Quentin J Leclerc, Audrey Duval, Didier Guillemot, Lulla Opatowski, and Laura Temime. Using contact network dynamics to implement efficient interventions against pathogen spread in hospital settings: A modelling study. PLoS Medicine, 21(7):e1004433, 2024.

[51] Agence technique de l’information sur l’hospitalisation. Guide méthodologique de production des informations relatives à l’activité médicale et à sa facturation en médecine, chirurgie, obstétrique et odontologie. Agence technique de l’information sur l’hospitalisation (ATIH), January 2024. URL https://www.atih.sante.fr/sites/default/files/public/content/4709/guide_methodo_mco_2024_version_provisoire_1.pdf. Guide méthodologique PMSI– MCO.

[52] Santé publique France. e-sin: signalement externe des infections nosocomiales. Santé publique France, 2024. URL https://www.santepubliquefrance.fr/e-sin-signalement-externe-des-infections-nosocomiales. National electronic reporting system for healthcare-associated infections.

[53] Narimane Nekkab, Pascal Crépey, Pascal Astagneau, Lulla Opatowski, and Laura Temime. Assessing the role of inter-facility patient transfer in the spread of carbapenemase-producing enterobacteriaceae: the case of france between 2012 and 2015. Scientific Reports, 10(1): 14910, 2020.

[54] Erin O’Fallon, Aurora Pop-Vicas, and Erika D’Agata. The emerging threat of multidrug-resistant gram-negative organisms in long-term care facilities. Journals of Gerontology Series A: Biomedical Sciences and Medical Sciences, 64(1):138–141, 2009.

[55] Ching Jou Lim, Allen C Cheng, Jacqueline Kennon, Denis Spelman, Dayna Hale, Gabrielle Melican, Hanna E Sidjabat, David L Paterson, David CM Kong, and Anton Y Peleg. Prevalence of multidrug-resistant organisms and risk factors for carriage in long-term care facilities: a nested case–control study. Journal of Antimicrobial Chemotherapy, 69(7): 1972–1980, 2014.

[56] Vikas Gupta, Michael J Satlin, C Yu Kalvin, Yehoda Martei, Lillian Sung, Lars F Westblade, Scott Howard, ChinEn Ai, and Diane C Flayhart. Incidence and prevalence of antimicrobial resistance in outpatients with cancer: a multicentre, retrospective, cohort study. The Lancet Oncology, 26(5):620–628, 2025.

[57] Mélanie Colomb-Cotinat, Amélie Jouzeau, Gaëlle Pedrono, Aurélie Chabaud, Christian Martin, Isabelle Poujol, Sylvie Maugat, Lory Dugravot, Catherine Dumartin, Anne Berger- Carbonne, et al. Estimating the number and incidence of carbapenemase-producing enterobacterales infections in france in 2020: A capture-recapture study. Infectious Diseases Now, 55(1):105016, 2025.

[58] Annie Herbert, Linda Wijlaars, Ania Zylbersztejn, David Cromwell, and Pia Hardelid. Data resource profile: hospital episode statistics admitted patient care (hes apc). International journal of epidemiology, 46(4):1093–1093i, 2017.

[59] Yin Mo, Anastasia Hernandez-Koutoucheva, Patrick Musicha, Denis Bertrand, David Lye, Oon Tek Ng, Shannon N Fenlon, Swaine L Chen, Moi Lin Ling, Wen Ying Tang, et al. Duration of carbapenemase-producing enterobacteriaceae carriage in hospital patients. Emerging infectious diseases, 26(9):2182, 2020.

[60] Steven G. Johnson. The NLopt Nonlinear-Optimization Package, 2008. URL https://github.com/stevengj/nlopt.

[61] Michael Hahsler, Matthew Piekenbrock, and Derek Doran. dbscan: Fast density-based clustering with R. Journal of Statistical Software, 91(1):1–30, 2019. doi: 10.18637/jss.v091.i01.

[62] Simon N Wood, Yannig Goude, and Simon Shaw. Generalized additive models for large data sets. Journal of the Royal Statistical Society Series C: Applied Statistics, 64(1):139–155, 2015.

[63] Santé publique France. Indice de défavorisation sociale (fdep) par commune. Odissé – Open Data des indicateurs de santé, 2026. URL https://odisse.santepubliquefrance.fr/explore/dataset/indice-de-defavorisation-sociale-fdep-par-commune/. FDep20 update carried out by Cnam using 2020 INSEE census and tax-income data; geography as of January 1, 2023. Last modified May 26, 2026.

[64] Julia M. Rohrer and Vincent Arel-Bundock. Models as prediction machines: How to convert confusing coefficients into clear quantities. Advances in Methods and Practices in Psychological Science, 9(2):25152459261424825, 2026. doi: 10.1177/25152459261424825.

[65] Waldo R Tobler. A computer movie simulating urban growth in the detroit region. Economic geography, 46(sup1):234–240, 1970.

[66] Andrew D. Cliff and J. Keith Ord. Spatial Processes: Models & Applications. Pion, London, 1981.

[67] Lance A. Waller and Carol A. Gotway. Applied Spatial Statistics for Public Health Data. Wiley, Hoboken, NJ, 2004.

[68] Roger Bivand and David W. S. Wong. Comparing implementations of global and local indicators of spatial association. TEST, 27(3):716–748, 2018. doi: 10.1007/s11749-018-0599-x.

[69] Luc Anselin. Local indicators of spatial association—lisa. Geographical analysis, 27(2): 93–115, 1995.

[70] Roger S. Bivand, Edzer Pebesma, and Virgilio Gómez-Rubio. Applied Spatial Data Analysis with R. Springer, New York, 2 edition, 2013. doi: 10.1007/978-1-4614-7618-4.

[71] Luc Anselin. The moran scatterplot as an esda tool to assess local instability in spatial association. In Spatial analytical perspectives on GIS, pages 111–126. Routledge, 2019.

[72] Yoav Benjamini and Yosef Hochberg. Controlling the false discovery rate: A practical and powerful approach to multiple testing. Journal of the Royal Statistical Society, Series B, 57 (1):289–300, 1995.

[73] Marcia Caldas de Castro and Burton H. Singer. Controlling the false discovery rate: A new application to account for multiple and dependent tests in local statistics of spatial association. Geographical Analysis, 38(2):180–208, 2006. doi: 10.1111/j.0016-7363.2006.00682.x.

[74] Vincent Arel-Bundock, Noah Greifer, and Andrew Heiss. How to interpret statistical models using marginaleffects for R and Python. Journal of Statistical Software, 111(9):1–32, 2024. doi: 10.18637/jss.v111.i09.

[75] ATIH. Guide méthodologique de production des informations relatives à l’activité médicale et à sa facturation en médecine, chirurgie, obstétrique et odontologie (pmsi-mco). Technical report, Agence Technique de l’Information sur l’Hospitalisation (ATIH), 2024. URL https://www.fhpmco.fr/wp-content/uploads/2024/02/guide_methodo_mco_2024_version_provisoire.pdf. Version provisoire.

[76] Anna D Broido and Aaron Clauset. Scale-free networks are rare. Nature communications, 10(1):1017, 2019.

